# The Efficacy of Psychological Interventions for Child and Adolescent PTSD: A Network Meta-Analysis

**DOI:** 10.1101/2024.04.09.24305537

**Authors:** Thole H. Hoppen, Lena Wessarges, Marvin Jehn, Julian Mutz, Ahlke Kip, Pascal Schlechter, Richard Meiser-Stedman, Nexhmedin Morina

## Abstract

**Importance:** Pediatric post-traumatic stress disorder (PTSD) is a common and debilitating mental disorder. Yet, a comprehensive network meta-analysis examining the efficacy of psychological interventions is lacking.

**Objective:** To synthesize all available evidence on psychological interventions for pediatric PTSD in a comprehensive network meta-analysis.

**Data Sources:** PsycINFO, MEDLINE, Web of Science, and PTSDpubs were searched from inception to January 2^nd^ 2024 and 74 related systematic reviews were screened.

**Study Selection:** Two independent raters screened publications for eligibility. Inclusion criteria were: Randomized controlled trial (RCT) with ten or more patients per arm examining a psychological intervention for pediatric PTSD compared to a control group in children and adolescents (< 19 years) with full or subthreshold PTSD.

**Data Extraction and Synthesis:** PRISMA guidelines were followed to synthesize and present evidence. Two independent raters extracted data and assessed risk of bias with Cochrane criteria. Random effects network meta-analyses were run.

**Main Outcome and Measures:** **S**tandardized mean differences (Hedges’ *g*) in PTSD severity.

**Results:** In total, 70 RCTs (N = 5,528 patients) were included. Most RCTs (74%) examined trauma-focused cognitive behavior therapy interventions (TF-CBTs). At treatment endpoint, TF-CBTs, EMDR, multi-disciplinary treatments (MDTs), and non-trauma-focused interventions (non-trauma-focused interventions) were all efficacious in reducing PTSD when compared to passive control conditions, with large pooled effects (*gs* ≥ 0.86, all *ps* < .001) in the random effects network meta-analysis. TF-CBTs produced the strongest short-term effects relative to both passive and active control conditions and across all sensitivity analyses. In a sensitivity analysis including only trials with parent involvement, TF-CBTs were significantly more efficacious in reducing PTSD than non-trauma-focused interventions (*g* = 0.35, *p* = .026). Results for mid-term (up to 5 months posttreatment) and long-term data (6-24 months posttreatment) were very similar.

**Conclusions and Relevance:** The present network meta-analysis is the most comprehensive summary of psychological treatments for pediatric PTSD to this date. Results confirm that TF-CBTs can efficaciously reduce PTSD symptom severity in children and adolescents in the short-, mid-, and long-term. More long-term data are needed for EMDR, MDTs, and non-trauma-focused interventions. Results of TF-CBTs are encouraging and disseminating these results may help reduce common treatment barriers.

**Key Points:** *Question:* How efficacious are psychological treatments for pediatric PTSD?

*Findings:* Trauma-focused cognitive behavior therapies (TF-CBTs) are currently the most evaluated treatment for pediatric PTSD (74% of included studies). Data for other interventions are emerging. At short-term, TF-CBTs, Eye Movement Desensitization and Reprocessing (EMDR), non-trauma-focused interventions, and multi-disciplinary treatments (MDTs) all significantly reduced pediatric PTSD relative to no treatment. TF-CBTs produced the strongest short- and long-term treatment effects. EMDR and MDTs had insufficient long-term data.

*Meaning:* TF-CBTs should be the first-line treatment recommendation for pediatric PTSD. While data for other treatment approaches emerged with some promising findings, more data (including long-term data) are needed to draw firmer conclusions.

---

One to two-thirds of children and adolescents from the general population report exposure to at least one traumatic event in their lifetimes.^1–4^ While most children and adolescents react resiliently to trauma, about one-fifth develop post-traumatic stress disorder (i.e., pediatric PTSD).^4,5^ Pediatric PTSD is a common, impairing,^4^ and often chronic^6^ mental disorder characterized by vivid re-experiencing of trauma, avoidance of trauma-related stimuli, changes in cognitions and emotions, and hyperarousal symptoms.^7^ Given the high prevalence and disease burden of pediatric PTSD,^8–10^ the examination and implementation of efficacious treatments constitutes a public health priority.

International treatment guidelines recommend cognitive behavior therapies with a trauma focus (TF-CBTs, e.g., TF-CBT^11^, prolonged exposure^12^, cognitive processing therapy^13^) as first-line treatment for pediatric PTSD.^14–19^ Research on other psychological interventions such as eye movement desensitization and reprocessing (EMDR) or non-trauma-focused interventions is also emerging. In recent years, number of published RCTs has increased substantially. To inform clinical practice about the relative efficacy of all treatment approaches, a comprehensive network meta-analysis (NMA) is required.

NMAs integrate data from both direct (i.e., comparison of arms within a given RCT) and indirect comparisons (i.e., comparisons of arms across RCTs), which enables conclusions about relative effects of all interventions based on all data.^20^ Three NMAs of psychological interventions for pediatric PTSD have been published.^21–23^ However, four omissions in previous works need to be addressed. First, a comprehensive NMA is needed. Caro et al.^23^ only focused on pediatric PTSD relating to sexual abuse (rather than any trauma type). Mavranezouli et al.^22^ analyzed follow-up data only up to four months posttreatment and cannot discern long-term efficacy. Xiang et al.^21^ included data from 56 RCTs published until 2020, compared to 70 RCTs published until 2024 in the present work. Second, no previous NMA included a sensitivity analysis of low risk of bias evidence. Low quality evidence may bias results in quantitative synthesis.^24^ Third, no NMA has performed sensitivity analyses concerning delivery format (e.g., individual delivery only or treatments with parent/caregiver involvement only) and efficacy may differ by delivery formats. Fourth, efficacy might also differ by age group (i.e., children vs. adolescents), and no NMA has yet performed an age-based sensitivity analysis. To enhance our understanding of relative treatment efficacy of interventions, the present work addresses these four omissions.

## Methods

We followed Preferred Reporting Items for Systematic Reviews and Meta-analyses (PRISMA) 2015 guidelines.^25^ The systematic literature search, data extraction, and risk of bias assessment were carried out independently by at least two authors. Disagreements were discussed between at least three authors (THH/LW/AK/NM). To address missing data, we sent data requests to corresponding authors, with a reminder one month later. The objectives and methods of the present NMA were pre-registered in the PROSPERO database (ID: CRD42020206290). We defined the main research question (Population, Intervention, Comparison, Outcome, and Study; PICOS) as follows: In children and adolescents with full or subthreshold PTSD (P), how do psychological interventions (I), compared to passive control conditions, active control conditions, or amongst different categories of interventions (C), perform in terms of lowering PTSD symptom severity (O) in randomized controlled trials (S)?

## Identification and Selection of Studies

### Search Strategy

For the timespan from inception to April 21^st^ 2022, we relied on the literature search of our previous work^26^ including 57 eligible RCTs. We conducted a new search with identical search strategy on January 2^nd^ 2024 covering literature published between April 21^st^ 2022 and January 2^nd^ 2024. Appendix A outlines the full search string. Briefly, we searched four large bibliometric health sciences databases: PsycINFO, MEDLINE, Web of Science, and PTSDpubs with various search terms for PTSD (ptsd OR ptss OR post-traumatic stress OR posttraumatic stress) and treatment (trial* OR treatment* OR intervention*) in all-field searches. Consistent with our previous search, no restrictions were applied to languages or publication formats. We also screened 74 recently published related systematic reviews (Appendix B) and the reference lists of included trials.

### Eligibility Criteria

In line with our previous work,^26^ we included trials that met all of the following inclusion criteria: 1) RCT, 2) investigating the efficacy of a psychological intervention for pediatric PTSD compared to a control condition, 3) all participants had PTSD complaints (full or sub-threshold PTSD), 4) sample mean age < 19.0 years, and 5) outcome data reported for at least ten participants per arm.

### Quality Assessment

Risk of bias was independently assessed by two authors (THH/NM) based on eight dichotomous quality criteria reported by Cuijpers and colleagues.^24^ These eight criteria originated partly from the Cochrane Collaboration criteria^27^ and authoritative criteria for evidence-based psychological interventions.^28^ In the present study, RCTs were defined as high-quality trials (i.e., with low risk of bias) when fulfilling at least six of the eight quality criteria (for quality criteria and quality ratings per trial see Appendices C & D, respectively). Initial agreement between independent raters was good (91.35%).

### Data Extraction

Trial characteristics (e.g., treatment delivery format), sample characteristics (e.g., mean age), and PTSD outcome data were extracted independently by at least two authors (THH/LW/AK/NM). When applicable, intent-to-treat (ITT) data were prioritized over completer data. When applicable, outcome data from clinician-based measures were prioritized over self-report-based data.

### Categorization of Psychological Interventions and Control Conditions

The present study compared four categories of psychological interventions based on the number of trials available: 1) TF-CBTs (i.e., any CBT-based intervention with a trauma focus, such as TF-CBT,^11^ prolonged exposure,^12^ or cognitive processing therapy^13^), 2) EMDR, 3) non-trauma-focused interventions (i.e., any intervention without a trauma focus), and 4) multi-disciplinary treatments (MDTs, i.e., treatments that combine techniques from at least two of the aforementioned categories of interventions, such as intensive multimodal group program^29^). Other trauma-focused interventions (i.e., interventions with a trauma focus but neither belonging to TF-CBTs or EMDR, such as expressive supportive groups^30^) were planned as fifth category, but lacked evidence (kes < 4). In the present work, *k* denotes the number of RCTs, whereas *kes* denotes the number of direct comparisons, which differ due to multi-arm trials. Control conditions were divided into passive control conditions (e.g., wait-list control) and active control conditions (e.g., treatment-as-usual, for all categorizations see, Appendix E).

### Categorization of Assessment Timepoints

To evaluate the short, mid, and long-term efficacy and consistent with previous research,^31^ we distinguished between three periods: 1) *post-treatment* (i.e., short-term), which we defined as assessments at treatment endpoint, 2) *mid-term*, which we defined as assessments of up to or equal to five months after the treatment endpoint, and 3) *long-term*, which we defined as assessments longer than five months after the treatment endpoint. When several assessments fell into the mid-term and long-term categories, the longest assessment was chosen.

### Outcomes

The primary outcome of interest was the standardized mean differences (Hedges’ *g*)^32^ in PTSD symptom severity between comparator groups.

### Statistical Analysis

Random effects NMAs were conducted given that high heterogeneity in outcomes was expected.^32^ Level of statistical significance was set to p-values below 0.05 (two-sided) for all analyses, including Egger’s test. Analyses were performed in R (version 4.1.1)^33^ with the netmeta package.^34^ Effect sizes (Hedges’ *g*) were first calculated at the study-level^35^ and then pooled and compared between all comparison dyads in NMA.^32^ Following Cohen,^36^ *g* was interpreted as small (0.20), medium (0.50), and large effect size (0.80). We only included intervention categories with minimally sufficient evidence base (i.e., *kes* ≥ 4)^37^ to ensure that effects were also estimated via a sufficient number of direct comparisons.^38^

For the transitivity assumption, we examined whether the distribution of various trial and sample characteristics was similar across comparison dyads and performed sensitivity analyses. We analyzed inconsistency between direct and indirect evidence^38^ globally (in the network) and locally (per comparison dyad)^25,39^ with the net splitting method^40^ and inspected net heat plots^41^. We performed inconsistency-corrected analyses when applicable. We calculated outlier-adjusted NMAs whenever (≥ 1) outliers were detected (i.e., ≥ 3.3 standard deviations above or below the pooled g)^42^. To examine the influence of small-study effects, we performed Egger’s test^43^ and inspected comparison-adjusted funnel plots^44^ (i.e., comparisons of interventions to passive and active control conditions only). We calculated the /^2^ and -r^2^ statistics as estimates of overall heterogeneity,^45^ and Q_het_ and Q_inc_ as estimates of heterogeneity within and between comparison dyads.^46^ We also calculated surface under the cumulative ranking (SUCRA; 50,000 resamples), allowing for a ranking by efficacy. To visualize distribution of available evidence, we build network graphs. In addition to main NMAs (across all data), we performed four sensitivity NMAs: 1) only high-quality trials, 2) only trials delivering treatment(s) individually, 3) only trials involving parents/caregivers in treatment, and 4) only trials with sample mean age < 12 years (i.e., involving mainly children) as well as only trials with sample mean age ≥ 12 years (i.e., involving mainly adolescents).

## Results

### Study Selection Process

The new search wave covered 8,845 electronic records with ten additional eligible RCTs. Thus, a total of 70 independent RCTs were eligible for the present purposes. Figure 1 details the study synthesis process.

**Figure 1.**
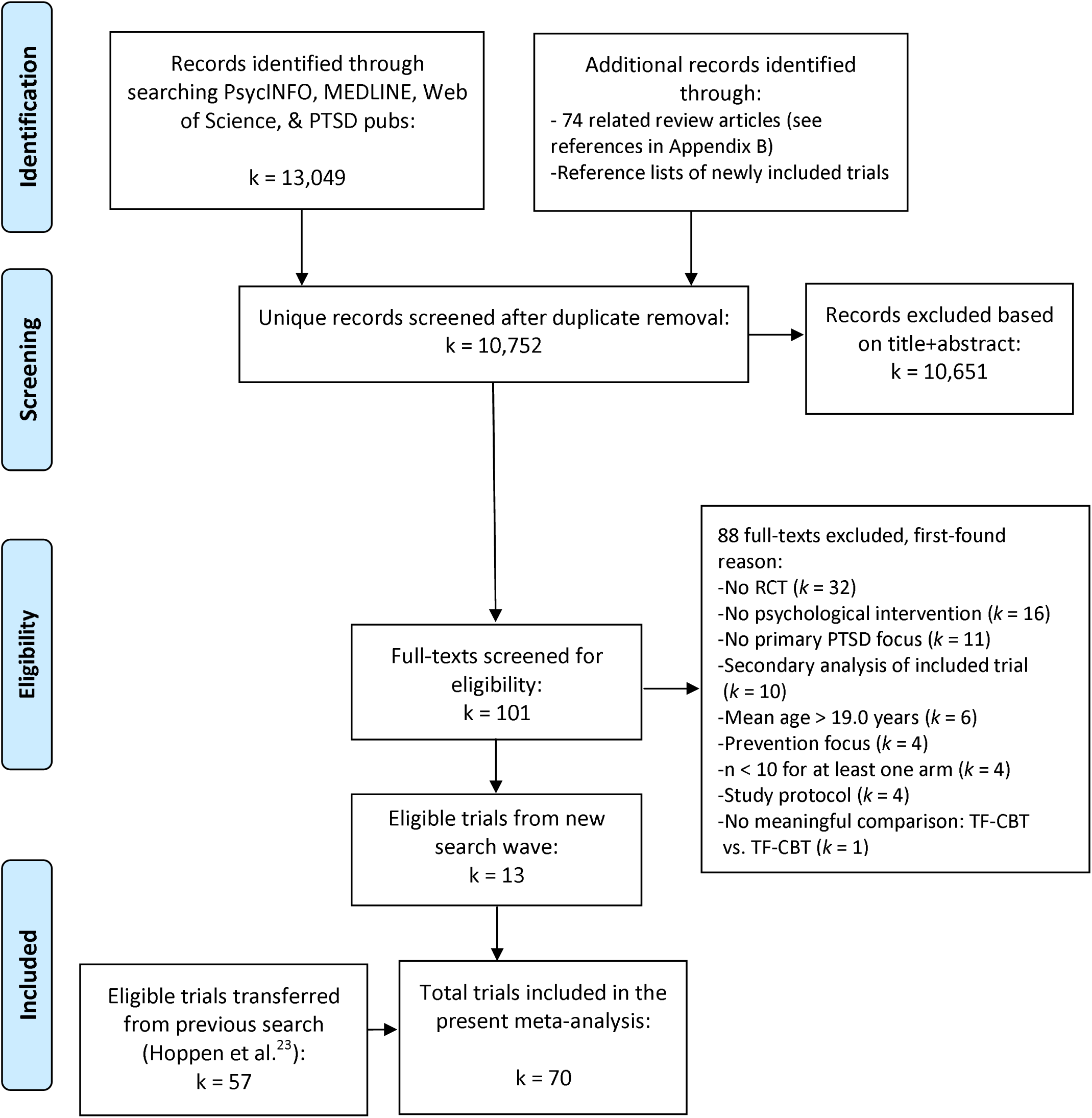
**PRISMA flow chart depicting the study selection process**

### Study Characteristics

The 70 RCTs reported data of 5,528 participants (for trial characteristics and their references, see Appendices F & G, respectively). Apart from one dissertation,^47^ all RCTs were peer-reviewed. Only Dorsey et al.^48^ reported on more than one RCT (i.e., four RCTs). In total, 41 RCTs (59% of trials) delivered interventions individually and 29 RCTs (41% of trials) involved parents or primary caregivers in treatments. Across trials reporting this information, mean number of total sessions was 10 and total duration of treatments (i.e., total sessions times length) was on average 11 hours (*SD* = 6.5 hours). In total, 40 RCTs (57% of trials) assessed follow-up data (range = 1mo-24mo posttreatment). ITT PTSD data were reported in 44 RCTs (63% of trials). In total, 41 RCTs (59%) were conducted in high-income countries and the remaining 29 RCTs (41%) in low and middle-income countries. However, two RCTs (conducted in the US and Germany) exclusively involved refugees originating from low-income countries.^49,50^ Across trials reporting this information (*k* = 52 or 74%), 90% of the participants met diagnostic criteria for PTSD at baseline. Fifty-seven trials (81%) involved mixed gender samples, whereas nine RCTs (13%) included only females and four RCTs (6%) only males. Across all trials, 60% of participants identified as females. Average age across trials was 12.21 years (*SD* = 3.08). In terms of trauma history, 31 RCTs (44%) included a sample with varying trauma histories. In the other trials, only participants with a particular trauma history were included, such as sexual assault (*k* = 10, 14%) or parental death (*k* = 6, 9%).

### Network Meta-analyses of Short, Mid, and Long-term Efficacy

#### Assumptions

Assumptions were mostly met. Apart from two analyses, no inconsistencies were observed. In the main NMA on mid-term efficacy, significant inconsistency (i.e., between direct and indirect evidence) was found for MDTs and a corrected analysis (i.e., without MDTs) was performed. In the sensitivity NMA on mid-term efficacy of treatments with individual delivery only, significant inconsistencies were found for all interventions (precluding corrected re-analysis) and results are thus not reported. The distribution of sample and methodological characteristics across comparison dyads is presented in Appendix H.

#### Network Graphs

Figure 2 shows the network graphs for the NMAs on short-, mid-, and long-term efficacy. Most available trials assessed TF-CBTs. Only TF-CBTs had enough accumulated evidence to be analyzed across all three timepoints.

**Figure 2.**
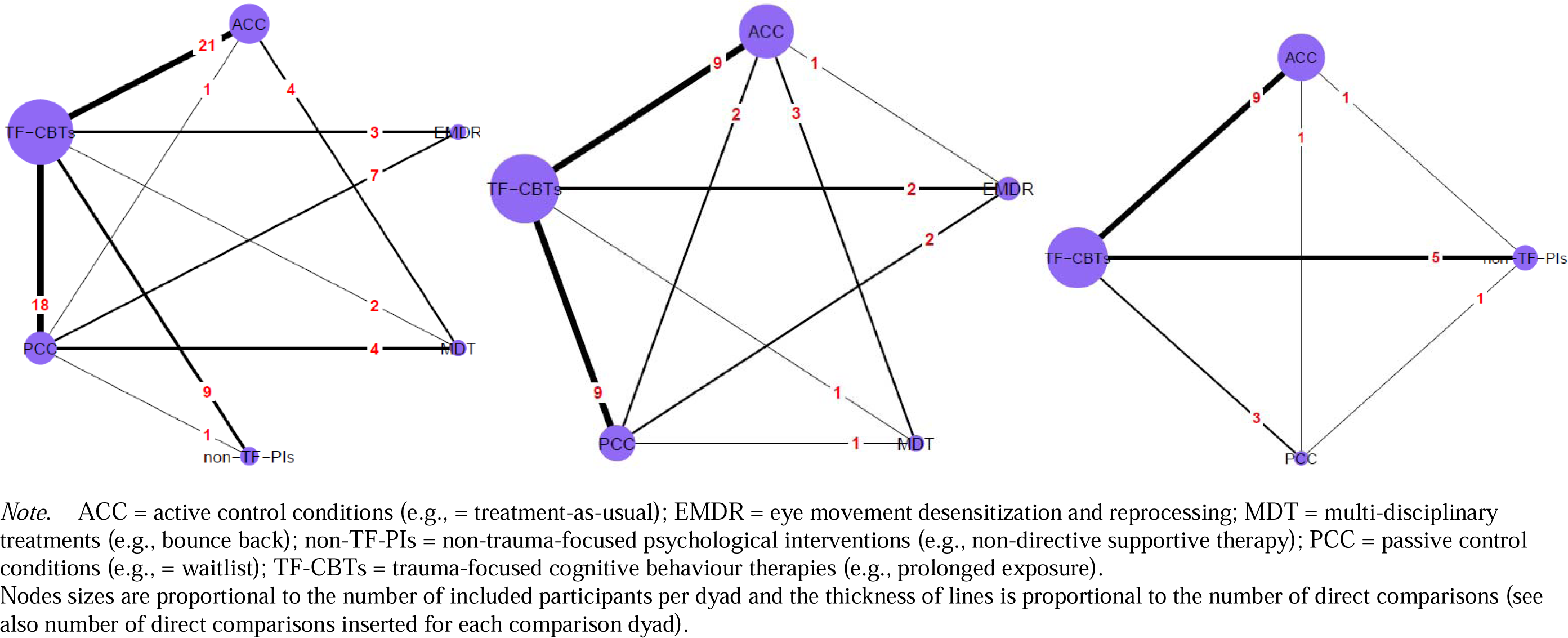
**Network graphs for the main analyses concerning short-term (left), mid-term (middle), and long-term (right) efficacy**

#### Network Meta-analysis of Short-term Efficacy

Table 1 provides all short-term results including sensitivity analyses. At treatment endpoint, all therapy categories – TF-CBTs, EMDR, MDTs, and non-trauma-focused interventions – were efficacious in treating pediatric PTSD compared to passive control conditions, with *g*s ranging from 0.86 for non-trauma-focused interventions to 1.06 for TF-CBTs (see Appendix I for the corresponding forest plot). Compared to active control conditions, only TF-CBTs (*g* = 0.55, *p* < .001) and MDTs (*g* = 0.43, *p* = .013) were efficacious (Appendix J). Differences in efficacy between treatment categories were not significant, with few or no direct comparisons for most comparison dyads. Heterogeneity in this main analysis of the short-term efficacy was large within and between comparison dyads (-r^2^ = 0.14, /^2^ = 68.9%; Q_total_ = 196.06, *df* = 61, p < .001; Q _et_ = 173.76, *df* = 50, p < .001; Q_inc_ = 21.40, *df* = 11, p = .030). No significant inconsistencies were detected in the net splitting method (Appendices K & L). No evidence for small-study effects was found (Appendix M). Two outliers^51,52^ investigating TF-CBTs were detected. Outlier-adjusted analysis excluding these trials produced similar results (Appendix N).

**Table 1.**
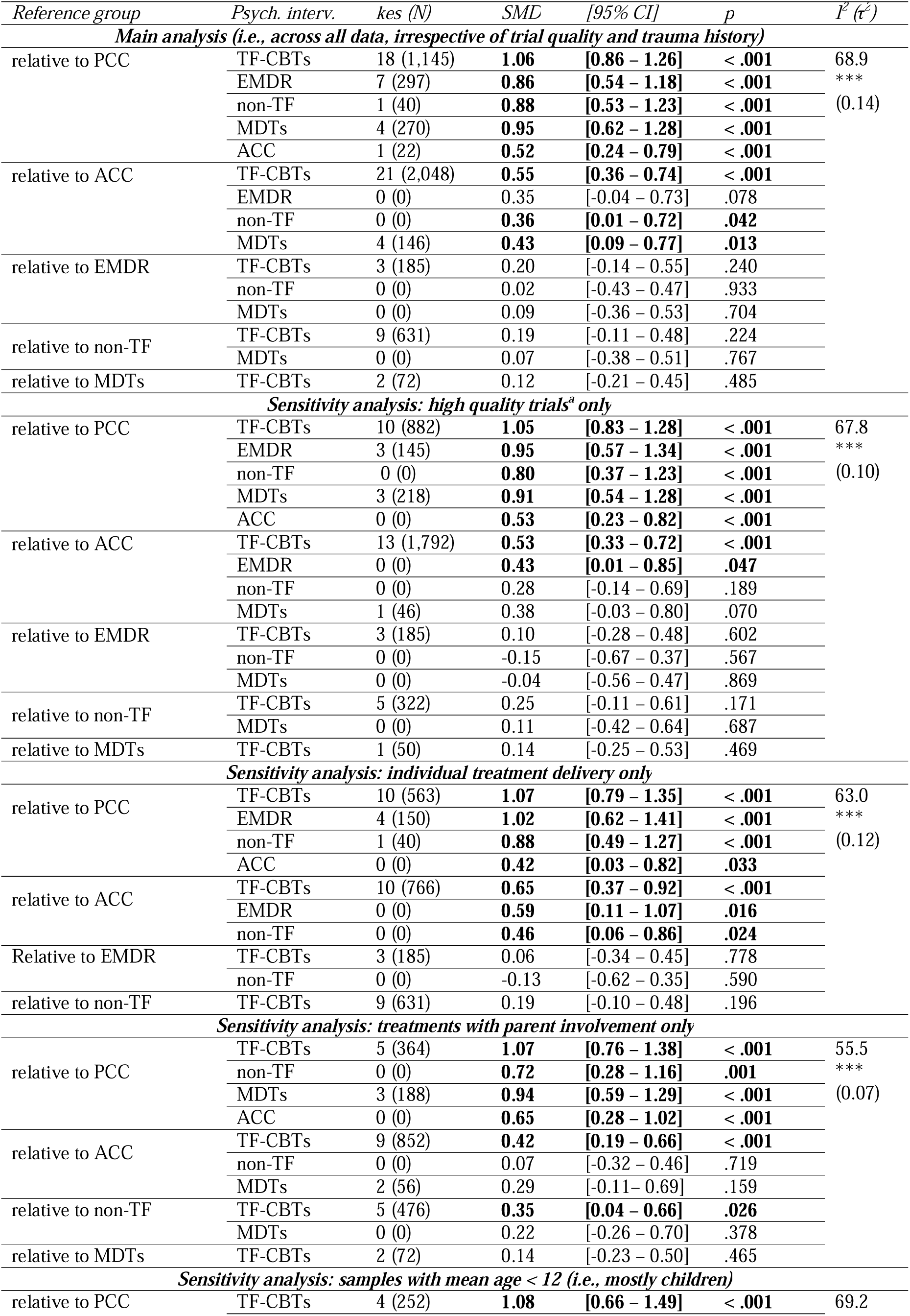
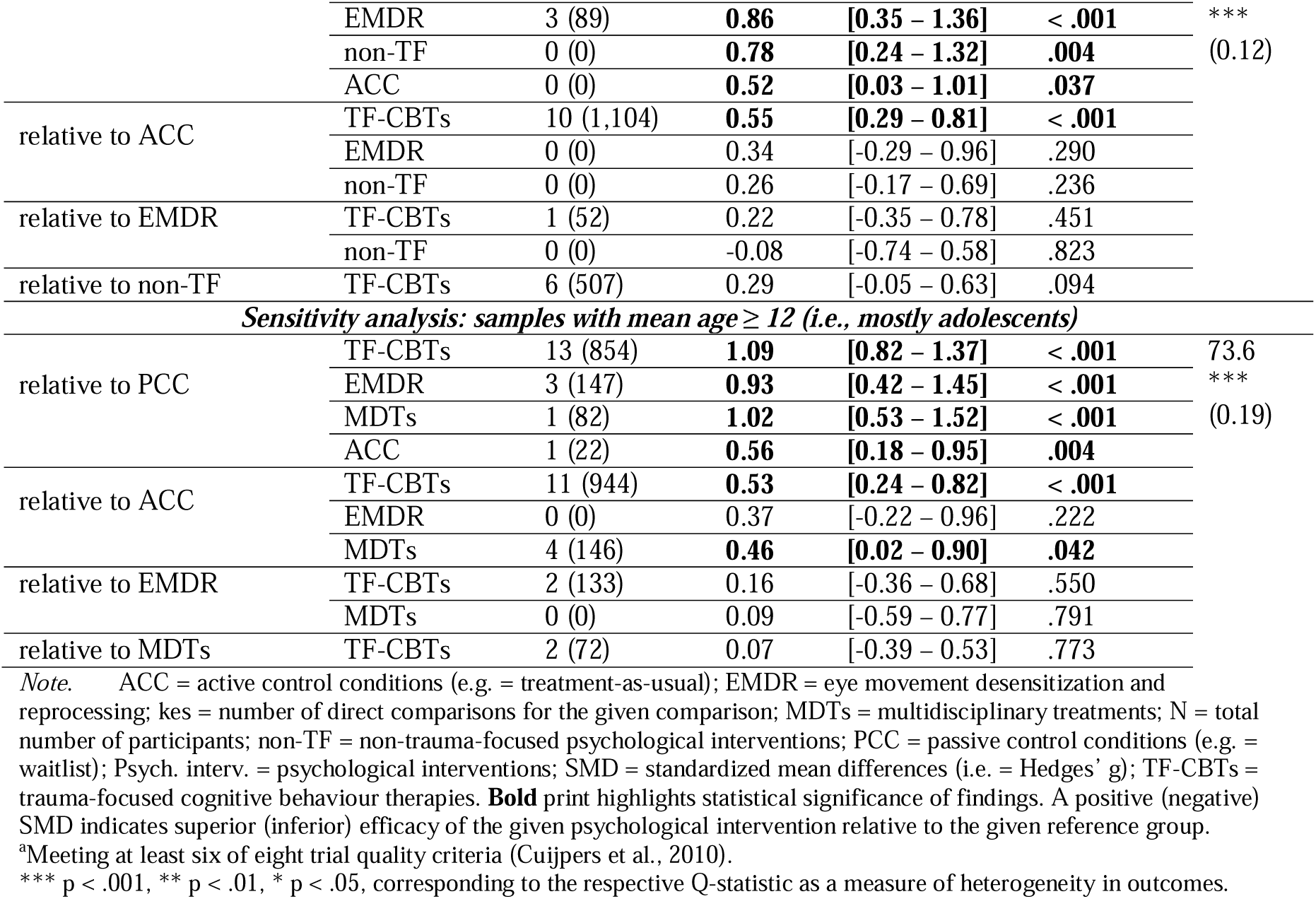
Short-term efficacy of psychological interventions for pediatric PTSD.

### Sensitivity Analyses for Short-term Efficacy

In high-quality trials only, the results for comparisons to passive control conditions were similar, with effect sizes ranging from 0.80 for non-trauma-focused interventions to 1.05 for TF-CBTs. Only TF-CBTs (*g* = 0.53, *p* < .001) and EMDR (*g* = 0.43, *p* = .047) produced significant short-term effects compared to active control conditions. In the sensitivity analysis concerning trials with individual treatment delivery only, results were similar to the main analysis with most favorable outcomes for TF-CBT. In the sensitivity analysis concerning only trials with parent/caregiver involvement, results were also similar for the comparison to passive control conditions, with all categories with sufficient evidence for synthesis (TF-CBT, MDTs, & non-trauma-focused interventions) producing significant effects. Compared to active control conditions, however, only TF-CBTs (*g* = 0.42, *p* < .001) produced a significant effect. Moreover, TF-CBTs with parent/caregiver involvement were more efficacious than non-trauma-focused interventions with caregiver involvement (*g* = 0.35, *p* = .026). In the sensitivity analysis of samples with mean age < 12 (mostly children), results were very similar with all intervention categories with sufficient evidence (TF-CBTs, EMDR, & non-trauma-focused interventions) yielding significant reductions in PTSD compared with passive controls (*gs* ≥ 0.78). Yet, only TF-CBTs produced a significant effect compared to active controls (*g* = 0.55, *p* < .001). In the sensitivity analysis of samples with mean age ≥ 12 (mostly adolescents), results were also very similar with TF-CBTs, EMDR, and MDTs yielding significant reductions in PTSD compared with passive controls (*gs* ≥ 0.93). Only TF-CBTs (*g* = 0.53, *p* < .001) and MDTs (*g* = 0.46, *p* = .042) produced significant effects compared to active controls.

#### Network Meta-analyses of Mid and Long-term Efficacy

Table 2 provides all results. For non-trauma-focused interventions, too few direct comparisons were available. At mid-term (up to 5 months posttreatment), TF-CBTs, EMDR, and MDTs were efficacious compared to passive control conditions with *gs* ranging from 0.59 for MDTs to 0.95 for EMDR (Appendix O). Compared to active control conditions, only EMDR (*g* = 0.52, *p* = .032) and TF-CBTs (*g* = 0.45, *p* = .002) produced significant mid-term effects (Appendix P). Heterogeneity in this main analysis concerning mid-term efficacy was large within and between comparison dyads (-r^2^ = 0.15, /^2^ = 66.4%; Q_total_ = 68.49, *df* = 23, p < .001; Q_□et_ = 49.40, *df* = 16, p < .001; Q_inc_ = 19.85, df = 7, p = .006). There was no evidence for small-study effects (Appendix Q). Significant inconsistency was detected for MDTs in the net splitting method (Appendices R & S). Results remained similar to those of the main analysis in a re-analysis excluding MDTs (Appendix T).

**Table 2.**
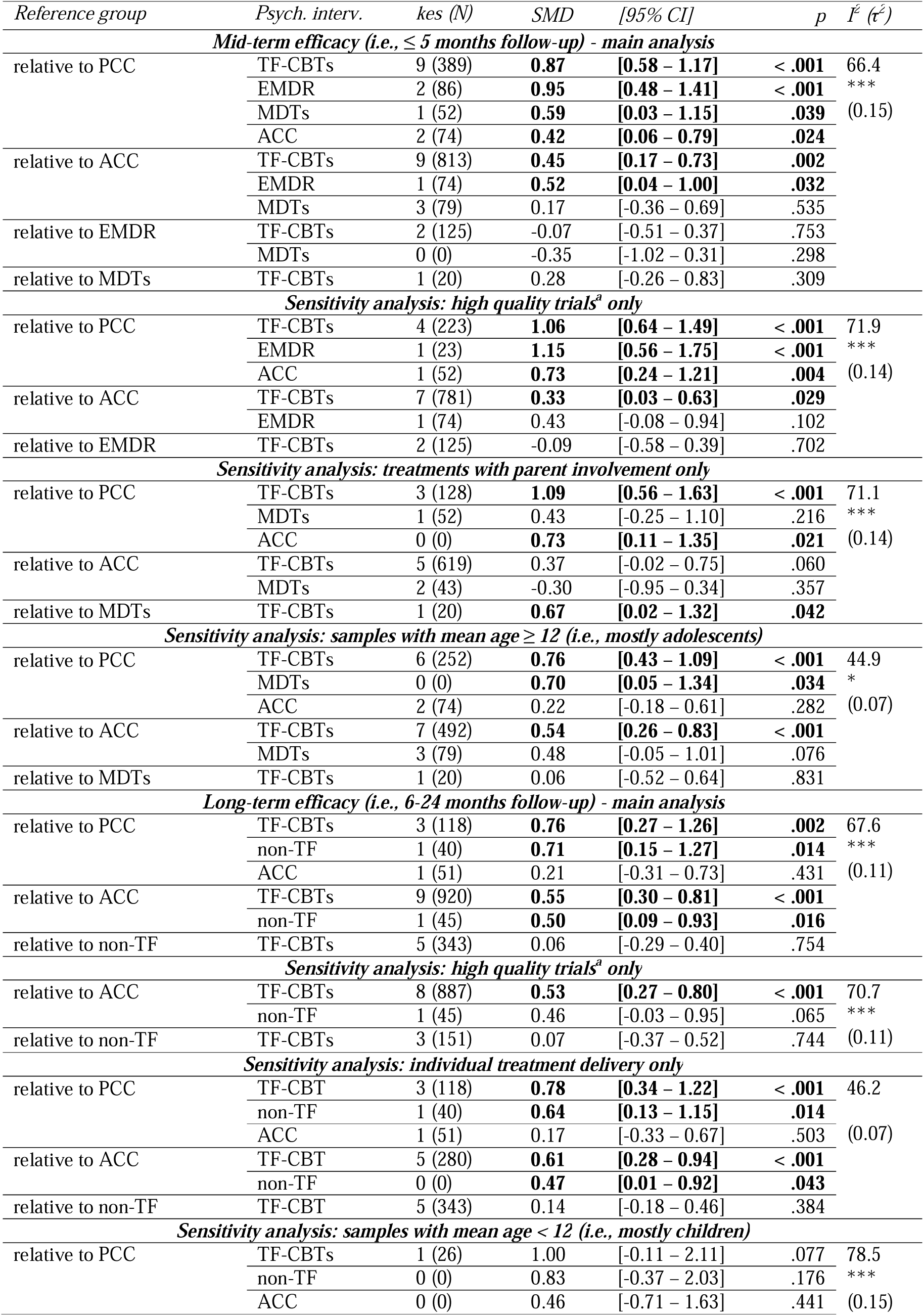
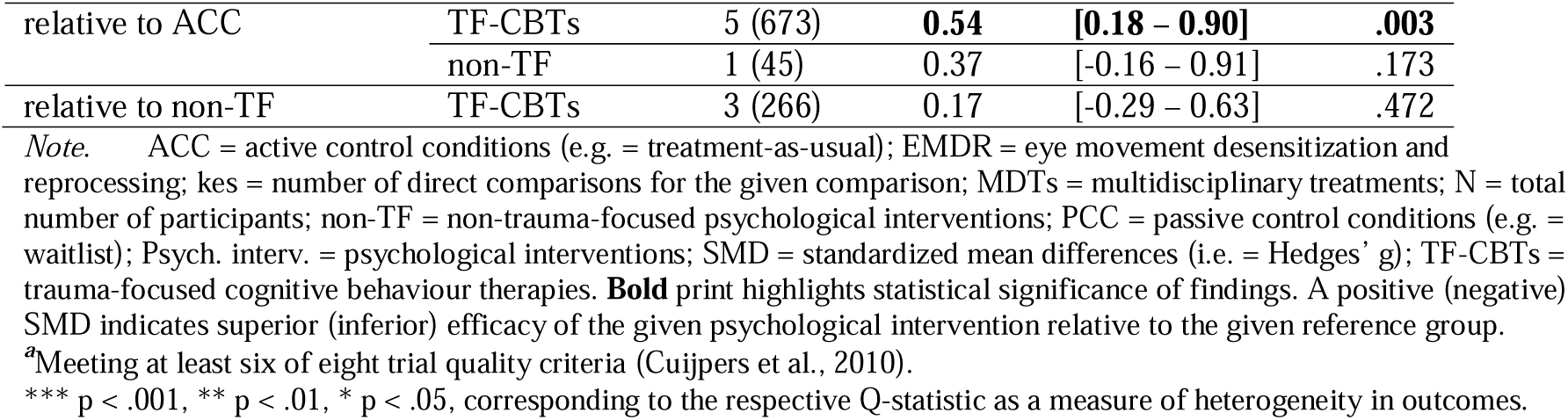
Mid-term efficacy (top) and long-term efficacy (bottom) of psychological interventions for pediatric PTSD.

| Reference group | Psych. interv. | kes (N) | SMD | [95% CI] | p | I<sup>2</sup> (τ<sup>2</sup>) |
| --- | --- | --- | --- | --- | --- | --- |
| **Mid-term efficacy (i.e., ≤ 5 months follow-up) - main analysis** |  |  |  |  |  |  |
| relative to PCC | TF-CBTs | 9 (389) | **0.87** | **[0.58 – 1.17]** | **< .001** | 66.4<br>***<br>(0.15) |
|  | EMDR | 2 (86) | **0.95** | **[0.48 – 1.41]** | **< .001** |  |
|  | MDTs | 1 (52) | **0.59** | **[0.03 – 1.15]** | **.039** |  |
|  | ACC | 2 (74) | **0.42** | **[0.06 – 0.79]** | **.024** |  |
| relative to ACC | TF-CBTs | 9 (813) | **0.45** | **[0.17 – 0.73]** | **.002** |  |
|  | EMDR | 1 (74) | **0.52** | **[0.04 – 1.00]** | **.032** |  |
|  | MDTs | 3 (79) | 0.17 | [-0.36 – 0.69] | .535 |  |
| relative to EMDR | TF-CBTs | 2 (125) | -0.07 | [-0.51 – 0.37] | .753 |  |
|  | MDTs | 0 (0) | -0.35 | [-1.02 – 0.31] | .298 |  |
| relative to MDTs | TF-CBTs | 1 (20) | 0.28 | [-0.26 – 0.83] | .309 |  |
| **Sensitivity analysis: high quality trials<sup>a</sup> only** |  |  |  |  |  |  |
| relative to PCC | TF-CBTs | 4 (223) | **1.06** | **[0.64 – 1.49]** | **< .001** | 71.9<br>***<br>(0.14) |
|  | EMDR | 1 (23) | **1.15** | **[0.56 – 1.75]** | **< .001** |  |
|  | ACC | 1 (52) | **0.73** | **[0.24 – 1.21]** | **.004** |  |
| relative to ACC | TF-CBTs | 7 (781) | **0.33** | **[0.03 – 0.63]** | **.029** |  |
|  | EMDR | 1 (74) | 0.43 | [-0.08 – 0.94] | .102 |  |
| relative to EMDR | TF-CBTs | 2 (125) | -0.09 | [-0.58 – 0.39] | .702 |  |
| **Sensitivity analysis: treatments with parent involvement only** |  |  |  |  |  |  |
| relative to PCC | TF-CBTs | 3 (128) | **1.09** | **[0.56 – 1.63]** | **< .001** | 71.1<br>***<br>(0.14) |
|  | MDTs | 1 (52) | 0.43 | [-0.25 – 1.10] | .216 |  |
|  | ACC | 0 (0) | **0.73** | **[0.11 – 1.35]** | **.021** |  |
| relative to ACC | TF-CBTs | 5 (619) | 0.37 | [-0.02 – 0.75] | .060 |  |
|  | MDTs | 2 (43) | -0.30 | [-0.95 – 0.34] | .357 |  |
| relative to MDTs | TF-CBTs | 1 (20) | **0.67** | **[0.02 – 1.32]** | **.042** |  |
| **Sensitivity analysis: samples with mean age ≥ 12 (i.e., mostly adolescents)** |  |  |  |  |  |  |
| relative to PCC | TF-CBTs | 6 (252) | **0.76** | **[0.43 – 1.09]** | **< .001** | 44.9<br>*<br>(0.07) |
|  | MDTs | 0 (0) | **0.70** | **[0.05 – 1.34]** | **.034** |  |
|  | ACC | 2 (74) | 0.22 | [-0.18 – 0.61] | .282 |  |
| relative to ACC | TF-CBTs | 7 (492) | **0.54** | **[0.26 – 0.83]** | **< .001** |  |
|  | MDTs | 3 (79) | 0.48 | [-0.05 – 1.01] | .076 |  |
| relative to MDTs | TF-CBTs | 1 (20) | 0.06 | [-0.52 – 0.64] | .831 |  |
| **Long-term efficacy (i.e., 6-24 months follow-up) - main analysis** |  |  |  |  |  |  |
| relative to PCC | TF-CBTs | 3 (118) | **0.76** | **[0.27 – 1.26]** | **.002** | 67.6<br>***<br>(0.11) |
|  | non-TF | 1 (40) | **0.71** | **[0.15 – 1.27]** | **.014** |  |
|  | ACC | 1 (51) | 0.21 | [-0.31 – 0.73] | .431 |  |
| relative to ACC | TF-CBTs | 9 (920) | **0.55** | **[0.30 – 0.81]** | **< .001** |  |
|  | non-TF | 1 (45) | **0.50** | **[0.09 – 0.93]** | **.016** |  |
| relative to non-TF | TF-CBTs | 5 (343) | 0.06 | [-0.29 – 0.40] | .754 |  |
| **Sensitivity analysis: high quality trials<sup>a</sup> only** |  |  |  |  |  |  |
| relative to ACC | TF-CBTs | 8 (887) | **0.53** | **[0.27 – 0.80]** | **< .001** | 70.7<br>***<br>(0.11) |
|  | non-TF | 1 (45) | 0.46 | [-0.03 – 0.95] | .065 |  |
| relative to non-TF | TF-CBTs | 3 (151) | 0.07 | [-0.37 – 0.52] | .744 |  |
| **Sensitivity analysis: individual treatment delivery only** |  |  |  |  |  |  |
| relative to PCC | TF-CBT | 3 (118) | **0.78** | **[0.34 – 1.22]** | **< .001** | 46.2<br>(0.07) |
|  | non-TF | 1 (40) | **0.64** | **[0.13 – 1.15]** | **.014** |  |
|  | ACC | 1 (51) | 0.17 | [-0.33 – 0.67] | .503 |  |
| relative to ACC | TF-CBT | 5 (280) | **0.61** | **[0.28 – 0.94]** | **< .001** |  |
|  | non-TF | 0 (0) | **0.47** | **[0.01 – 0.92]** | **.043** |  |
| relative to non-TF | TF-CBT | 5 (343) | 0.14 | [-0.18 – 0.46] | .384 |  |
| **Sensitivity analysis: samples with mean age < 12 (i.e., mostly children)** |  |  |  |  |  |  |
| relative to PCC | TF-CBTs | 1 (26) | 1.00 | [-0.11 – 2.11] | .077 | 78.5<br>***<br>(0.15) |
|  | non-TF | 0 (0) | 0.83 | [-0.37 – 2.03] | .176 |  |
|  | ACC | 0 (0) | 0.46 | [-0.71 – 1.63] | .441 |  |

|  |  |  |  |  |  |
| --- | --- | --- | --- | --- | --- |
| relative to ACC | TF-CBTs | 5 (673) | **0.54** | **[0.18 – 0.90]** | **.003** |
|  | non-TF | 1 (45) | 0.37 | [-0.16 – 0.91] | .173 |
| relative to non-TF | TF-CBTs | 3 (266) | 0.17 | [-0.29 – 0.63] | .472 |

*Note.* ACC = active control conditions (e.g. = treatment-as-usual); EMDR = eye movement desensitization and reprocessing; kes = number of direct comparisons for the given comparison; MDTs = multidisciplinary treatments; N = total number of participants; non-TF = non-trauma-focused psychological interventions; PCC = passive control conditions (e.g. = waitlist); Psych. interv. = psychological interventions; SMD = standardized mean differences (i.e. = Hedges' g); TF-CBTs = trauma-focused cognitive behaviour therapies. **Bold** print highlights statistical significance of findings. A positive (negative) SMD indicates superior (inferior) efficacy of the given psychological intervention relative to the given reference group.
<sup>a</sup>Meeting at least six of eight trial quality criteria (Cuijpers et al., 2010).
\*\*\* p < .001, \*\* p < .01, \* p < .05, corresponding to the respective Q-statistic as a measure of heterogeneity in outcomes.

In the long-term (6-to-24 months posttreatment), only TF-CBTs and non-trauma-focused interventions had sufficient evidence for synthesis. Compared to passive control conditions, both TF-CBTs (*g* = 0.76, *p* = .002) and non-trauma-focused interventions (*g* = 0.71, *p* = .014) produced significant effects (Appendix U). Both TF-CBTs (*g* = 0.55, *p* < .001) and non-trauma-focused interventions (*g* = 0.50, *p* = .016) were also efficacious when compared to active control conditions (Appendix V). Heterogeneity in this main analysis concerning long-term efficacy was large within and between comparison dyads (-r^2^ = 0.11, /^2^ = 67.6%; Q_total_ = 46.30, *df* = 15, p < .001; Q_□et_ = 32.33, *df* = 10, p < .001; Q_inc_ = 14.02, df = 5, p = .016). No inconsistencies (Appendices W & X) and no evidence for small-study effects (Appendix Y) were found.

### Sensitivity Analyses for Mid- and Long-term Efficacy

At mid-term, sensitivity analysis on high-quality trials could be conducted with TF-CBTs and EMDR only. EMDR (*g* = 1.15, *p* < .001) and TF-CBTs (*g* = 1.06, *p* < .001) produced large treatment effects compared to passive control conditions. Compared to active control conditions, however, only TF-CBTs (*g* = 0.33, *p* = .029) produced a significant treatment effect. The sensitivity analysis concerning trials with individual treatment delivery only was infeasible given detected inconsistency for all interventions. The sensitivity analysis of trials with parent/caregiver involvement produced similar results to the main analysis. Yet, TF-CBTs were found more efficacious than MDTs (*g* = 0.67, *p* = .042). The sensitivity analysis of trials with mean age below 12 was infeasible due to lacking evidence. The sensitivity analysis of trials with mean age ≥ 12 (mainly adolescents) produced similar results to the main analysis. TF-CBT (*g* = 0.76, *p* < .001) and MDTs (*g* = 0.70, *p* = .034) both produced significant effects relative to passive controls. Yet, only TF-CBT (*g* = 0.54, *p* < .001) was found significantly superior to active controls.

At long-term, only TF-CBTs and non-trauma-focused interventions had sufficient accumulated data. Passive control conditions were also lacking. Compared to active control conditions in high-quality trials, only TF-CBTs produced a significant moderate-sized treatment effect (*g* = 0.53, *p* < .001). Sensitivity analysis concerning trials with individual treatment delivery only produced similar results to the main analysis. Sensitivity analysis concerning trials with parent/caregiver involvement was infeasible due to lacking long-term evidence (kes < 4). The sensitivity analysis of trials with mean age ≥ 12 was infeasible due to lacking evidence. The sensitivity analysis of trials with mean age < 12 (mainly involving children) produced similar results to the main analysis, with only TF-CBT (but not non-trauma-focused interventions) producing significant effects compared to active control conditions (*g* = 0.54, *p* = .003).

### Ranking of Efficacy

Table 3 shows SUCRA rankings. TF-CBTs were the highest ranking category of interventions at all timepoints and all analyses, except at mid-term when it was second to EMDR.

**Table 3.** Efficacy rankings of psychological interventions for pediatric PTSD.

|  | SUCRA | SUCRA | SUCRA | SUCRA | SUCRA | SUCRA | SUCRA | SUCRA | SUCRA | SUCRA | SUCRA | SUCRA | SUCRA | SUCRA | SUCRA | SUCRA |
| --- | --- | --- | --- | --- | --- | --- | --- | --- | --- | --- | --- | --- | --- | --- | --- | --- |
|  | @short-term | @short-term | @short-term | @short-term | @short-term | @short-term | @short-term | @mid-term | @mid-term | @mid-term | @mid-term | @mid-term | @mid-term | @long-term | @long-term | @long-term |
|  | - all data (main analysis) | - outlier-adjusted | - high-quality trials only | - individual delivery of treatments only | - treatments with caregiver involvement only | - samples with mean age < 12 (mostly children) | - samples with mean age ≥ 12 (mostly adolescents) | - all data (main analysis) | - MDTs deleted due to inconsistency | - high-quality trials only | - treatments with caregiver involvement only | - samples with mean age ≥ 12 (mostly adolescents) | - all data (main analysis) | - high-quality trials only | - individual delivery of treatments only | - samples with mean age < 12 (mostly children) |
| TF-CBTs | 0.90 | 0.89 | 0.87 | 0.88 | 0.94 | 0.87 | 0.83 | 0.81 | 0.80 | 0.77 | 0.98 | 0.86 | 0.87 | 0.81 | 0.93 | 0.91 |
| EMDR | 0.58 | 0.61 | 0.71 | 0.77 | NA | 0.69 | 0.64 | 0.87 | 0.86 | 0.87 | NA | NA | NA | NA | NA | NA |
| MDTs | 0.70 | 0.72 | 0.66 | NA | 0.74 | NA | 0.74 | 0.50 | NA<sup>a</sup> | NA | 0.36 | 0.78 | NA | NA | NA | NA |
| Non-TF | 0.60 | 0.57 | 0.52 | 0.60 | 0.46 | 0.60 | NA | NA | NA | NA | NA | NA | 0.79 | 0.67 | 0.73 | 0.68 |
| ACC | 0.21 | 0.21 | 0.23 | 0.25 | 0.36 | 0.34 | 0.28 | 0.32 | 0.34 | 0.36 | 0.62 | 0.31 | 0.26 | 0.02<sup>b</sup> | 0.25 | 0.29 |
| PCC | 0.00 | 0.00 | 0.00 | 0.00 | 0.00 | 0.01 | 0.00 | 0.01 | 0.00 | 0.00 | 0.04 | 0.05 | 0.07 | NA | 0.09 | 0.12 |

*Note.* ACC = active control conditions (e.g. = treatment-as-usual); @long-term = long-term efficacy (i.e., assessments 6-24 months after treatment endpoint); @mid-term = mid-term efficacy (i.e., assessments ≤ 5 months after treatment endpoint); @short-term = short-term efficacy (i.e., assessments at treatment endpoint); EMDR = eye movement desensitization and reprocessing; high-quality trials only = sensitivity-analysis exclusively involving high-quality trials (i.e., trials fulfilling at least six of eight quality criteria); MDTs = multi-disciplinary treatments; NA = not applicable given insufficient accumulated evidence (i.e., $k < 4$ ); Non-TF = non-trauma-focused psychological interventions (i.e., psychological interventions without a trauma focus); PCC = passive control conditions (i.e., waitlist); SUCRA = surface under the cumulative ranking (i.e., efficacy ranked by means of surface under the cumulative ranking with 50,000 resamples); TF-CBTs = trauma-focused cognitive behaviour therapies.
<sup>a</sup>Excluded from this analysis given that significant inconsistency was detected.
<sup>b</sup>Reference group for this analysis given insufficient number of direct comparisons ( $k < 4$ ) for PCC.

## Discussion

The present work synthesized data from 70 RCTs in a comprehensive NMA. TF-CBTs are currently the most evaluated and the most efficacious category of treatments for PTSD in children and adolescents, followed by (in this order) EMDR, MDTs, and non-trauma-focused interventions. This supports recommendations of international treatment guidelines for pediatric PTSD, including the International Society for Traumatic Stress Studies^18^ and the National Institute of Clinical Excellence.^17^ Our review confirms and extends previous NMAs^21–23^ and pairwise meta-analyses.^26,53,54^ TF-CBTs were efficacious in treating pediatric PTSD relative to different comparators (passive and active control conditions), across time (short-, mid-, and long-term follow-up), and when restricting analyses to trials with high-quality, utilizing an individual delivered intervention, with parent involvement, and to samples involving mainly children or adolescents, respectively. These results are important for the training of therapist and treatment implementation in clinical practice and might help in reducing common treatment barriers.

Our review also reveals remaining gaps in the literature. While short and mid-term efficacy of EMDR was supported, our review highlights the lack of evidence concerning long-term follow-up data. The present results therefore support some international treatment guidelines^17,19^ that list EMDR as second-line treatment recommendation. Data for two other categories of interventions – MDTs and non-trauma-focused interventions – are emerging. More data (including long-term data), however, are needed to robustly investigate the (relative) efficacy of these two categories as well as EMDR. For the time being, only the efficacy of TF-CBTs can be robustly supported given that about three quarters of the available data concern TF-CBTs. There was some evidence for TF-CBTs producing significantly higher effects than non-trauma-focused interventions. Yet, more data is needed to draw firmer conclusions with regards to relative efficacy between categories of interventions.

### Limitations

Five limitations should be noted. First, the categories of non-trauma-focused interventions, MDTs, and other trauma-focused interventions are heterogenous with regards to theoretical foundations of included interventions. However, there is no solid ground for further sub-categorization given the low number of available RCTs. As more RCTs accumulate, more homogenous categorizations will become feasible. Second, we found evidence for inconsistency in the NMA regarding mid-term efficacy data. However, results remained similar in a consistency-corrected re-analysis. Third, our age group sensitivity analyses provide approximate estimations of treatment effects for children and adolescents, as the categorization was based on sample mean age. An individual patient data meta-analysis would allow for a solid differentiation of effects for children and adolescents, which was beyond the scope of the present work. Fourth, while the distinction between passive and active control conditions is a strength of the present work, active control condition such as treatment-as-usual can comprise very different elements, depending on the context. Future research might be able to disentangle this heterogenous comparator group in a more fine-grained way. Fifth, relatively low rates of reported ITT data are concerning and trialists are encouraged to report ITT data (including ITT means and SDs) in future trials.

### Conclusions

There is robust evidence that pediatric PTSD can be effectively treated by psychological treatments, in particular TF-CBTs. A large evidence base for TF-CBTs supports efficacy at short, mid, and long-term, relative to both passive and active control conditions and in high-quality trials. A comparably thin evidence base also supports the short and mid-term efficacy of EMDR and to a lesser extent MDTs and non-trauma-focused interventions. More high-quality trials including long-term assessments of PTSD are needed.

## Statements

### Author Contributions

THH, NM, LW, AK, MJ, and JM had full access to all of the data in the study and take responsibility for the integrity of the data and the accuracy of the data analysis.

Concept and design: THH and NM.

Acquisition, analysis, or interpretation of data: All authors.

Drafting of the manuscript: THH and RM.

Critical review of the manuscript for important intellectual content: All authors.

Statistical analysis: THH, MJ, LW, JM.

Obtained funding: Not applicable (no funding).

Administrative, technical, or material support: THH.

Supervision: THH and NM.

### Data Sharing Statement

The datasets and the R script are available on request via email to Dr. Thole H. Hoppen.

### Additional Contributions

We thank the authors of the included studies for their work and for responding to data request emails and the provision of data.

