## Appendix for "The Efficacy of Psychological Interventions for Child and Adolescent PTSD: A Network Meta-Analysis"

Short Title: Efficacy of psychological interventions for pediatric PTSD

*Corresponding Author

Thole H. Hoppen, Ph.D.

Institute of Psychology

University of Münster

Fliednerstr. 21

48149 Münster

Germany

Hyperlinked Contents

**Appendix A:**	**Search string used for systematic literature search**

**Appendix B:	References of screened reviews as part of the systematic literature search**

**Appendix C:	Quality criteria for risk of bias assessment**

**Appendix D:	Quality coding of included trials**

**Appendix E:	Categorization of interventions and control conditions**

**Appendix F:	Trial characteristics of included trials**

**Appendix G:	References of studies included in the present network meta-analysis**

**Appendix H: Trial and sample characteristics across comparison dyads**

**Appendix I:	Short-term efficacy: Forest plot compared to passive control conditions**

**Appendix J:	Short-term efficacy: Forest plot compared to active control conditions**

**Appendix K: Short-term efficacy: Net splitting results**

**Appendix L:	Short-term efficacy: Net heat plot**

**Appendix M:	Short-term efficacy: Funnel plot**

**Appendix N:	Short-term efficacy: Outlier-adjusted results**

**Appendix O:	Mid-term efficacy: Forest plot compared to passive control conditions**

**Appendix P:	Mid-term efficacy: Forest plot compared to active control conditions**

**Appendix Q:	Mid-term efficacy: Funnel plot**

**Appendix R: Mid-term efficacy: Net splitting results**

**Appendix S:	Mid-term efficacy: Net heat plot** **before (top) and after (bottom) exclusion of MDTs**

**Appendix T:	Mid-term efficacy: MDTs excluded due to detected inconsistency**

**Appendix U:	Long-term efficacy: Forest plot compared to passive control conditions**

**Appendix V:	Long-term efficacy: Forest plot compared to active control conditions**

**Appendix W: Long-term efficacy: Net splitting results**

**Appendix X:	Long-term efficacy: Net heat plot**

**Appendix Y:	Long-term efficacy: Funnel plot**

**Appendix A:** **Search string used for systematic literature search**

| Databases | Search Terms |
| --- | --- |
| MEDLINE and PsycINFO | ( TI ( ptsd OR ptss OR post-traumatic stress OR posttraumatic stress OR post-traumatic syndrome OR posttraumatic syndrome) OR AB ( ptsd OR ptss OR post-traumatic stress OR posttraumatic stress OR post-traumatic syndrome OR posttraumatic syndrome ) OR SU ( ptsd OR ptss OR post-traumatic stress OR posttraumatic stress OR post-traumatic syndrome OR posttraumatic syndrome ) ) AND ( TI ( treatment* OR intervention* OR therap* OR psychotherap* OR exposure OR counse*ing OR trial* ) OR AB ( treatment* OR intervention* OR therap* OR psychotherap* OR exposure OR counse*ing OR trial* ) OR SU ( treatment* OR intervention* OR therap* OR psychotherap* OR exposure OR counse*ing OR trial* ) ) |
| PTSDpubs | (ptsd OR ptss OR post-traumatic stress OR posttraumatic stress OR post-traumatic syndrome OR posttraumatic syndrome) AND (treatment* OR intervention* OR therap* OR psychotherap* OR exposure OR counse*ing OR trial*) |
| Web of Science | ALL=( ptsd OR ptss OR post-traumatic stress OR posttraumatic stress OR post-traumatic syndrome OR posttraumatic syndrome ) AND ALL=( treatment* OR intervention* OR therap* OR psychotherap* OR exposure OR counse*ing OR trial* ) |
| Note that the search string contains both APA thesaurus and MeSH search terms. | |

**Appendix B: References of screened reviews as part of the systematic literature search**

1. Alozkan‐Sever C, Uppendahl JR, Cuijpers P, et al. Research Review: Psychological and psychosocial interventions for children and adolescents with depression, anxiety, and post‐traumatic stress disorder in low‐and middle‐income countries–a systematic review and meta‐analysis. *J Child Psychol Psychiatry*. 2023;64(12):1776-1788. doi:10.1111/jcpp.13891.
2. Alzaghoul AF, McKinlay AR, Archer M. Post-traumatic stress disorder interventions for children and adolescents affected by war in low-and middle-income countries in the Middle East: systematic review. *BJPsych Open*. 2022;8(5):1-16. doi:10.1192/bjo.2022.552.
3. Annous N, Al-Hroub A, El Zein F. A systematic review of empirical evidence on art therapy with traumatized refugee children and youth. *Front Psychol*. 2022;13(5). doi:10.3389/fpsyg.2022.811515.
4. Badesha K, Wilde S, Dawson DL. Mental health mobile app use to manage psychological difficulties: An umbrella review. *Ment Health Rev (Brighton)*. 2022;27(3):241-280. doi:10.1108/MHRJ-02-2021-0014.
5. Baetz CL, Branson CE, Weinberger E, et al. The effectiveness of PTSD treatment for adolescents in the juvenile justice system: A systematic review. *Psychol Trauma*. 2022;14(4):642-652. doi:10.1037/tra0001073.
6. Bandelow B, Allgulander C, Baldwin DS, et al. World Federation of Societies of Biological Psychiatry (WFSBP) guidelines for treatment of anxiety, obsessive-compulsive and posttraumatic stress disorders–Version 3. Part I: Anxiety disorders. *World J Biol Psychiatry*. 2023;24(2):79-117. doi:10.1080/15622975.2022.2086295.
7. Braito I, Rudd T, Buyuktaskin D, Ahmed M, Glancy C, Mulligan A. Review: systematic review of effectiveness of art psychotherapy in children with mental health disorders. *Ir J Med Sci*. 2022;191(3):1369-1383. doi:10.1007/s11845-021-02688-y.
8. Byrne G. A systematic review of treatment interventions for individuals with intellectual disability and trauma symptoms: A review of the recent literature. *Trauma Violence Abuse*. 2022;23(2):541-554. doi:10.1177/1524838020960219.
9. Cantor AG, Nelson HD, Pappas M, et al. Telehealth for Women’s Preventive Services for Reproductive Health and Intimate Partner Violence: a Comparative Effectiveness Review. *J Gen Intern Med*. 2023;38(7):1735-1743. doi:10.1007/s11606-023-08033-6.
10. Caro P, Turner W, Caldwell DM, Macdonald G. Comparative effectiveness of psychological interventions for treating the psychological consequences of sexual abuse in children and adolescents: a network meta‐analysis. *Cochrane Database Syst Rev*. 2023;6:CD013361.
11. Capaldi JM, Shabanian J, Finster LB, et al. Post-traumatic stress symptoms, post-traumatic stress disorder, and post-traumatic growth among cancer survivors: a systematic scoping review of interventions. *Health Psychol Rev*. 2023:1-34. doi:10.1080/17437199.2022.2162947.
12. Cowling MM, Anderson JR. The effectiveness of therapeutic interventions on psychological distress in refugee children: A systematic review. *J Clin Psychol*. 2023;79(8):1857-1874. doi:10.1002/jclp.23479.
13. Davis RS, Meiser-Stedman R, Afzal N, et al. Systematic Review and Meta-analysis: Group-based interventions for treating posttraumatic stress symptoms in children and adolescents. *J Am Acad Child Adolesc Psychiatry*. 2023;62(11):1217-1232. doi:10.1016/j.jaac.2023.02.013.
14. de Haan A, Meiser-Stedman R, Landolt MA, et al. Efficacy and moderators of efficacy of cognitive behavioural therapies with a trauma focus in children and adolescents: an individual participant data meta-analysis of randomised trials. *Lancet Child Adolesc Health*. 2023;8(1):28-39.
15. Diano F, Sica LS, Ponticorvo M. A systematic review of mobile apps as an adjunct to psychological interventions for emotion dysregulation. *Int J Environ Res Public Health*. 2023;20(2). doi:10.3390/ijerph20021431.
16. English A, McKibben E, Sivaramakrishnan D, Hart N, Richards J, Kelly P. A Rapid Review Exploring the Role of Yoga in Healing Psychological Trauma. *Int J Environ Res Public Health*. 2022;19(23). doi:10.3390/ijerph192316180.
17. Genç E. The effectiveness of trauma treatment approaches in refugee children and adolescents: A systematic review. *Child Youth Serv Rev*. 2022;141.
18. Grainger L, Thompson Z, Morina N, Hoppen T, Meiser‐Stedman R. Associations between therapist factors and treatment efficacy in randomized controlled trials of trauma‐focused cognitive behavioral therapy for children and youth: A systematic review and meta‐analysis. *J Trauma Stress*. 2022;35(5):1405-1419.
19. Grande AJ, Hoffmann MS, Evans-Lacko S, et al. Efficacy of school-based interventions for mental health problems in children and adolescents in low and middle-income countries: A systematic review and meta-analysis. *Front Psychiatry*. 2023;13. doi:10.3389/fpsyt.2022.1012257.
20. Hermosilla S, Forthal S, Sadowska K, Magill EB, Watson P, Pike KM. We need to build the evidence: A systematic review of psychological first aid on mental health and well‐being. *J Trauma Stress*. 2023;36(1):5-16. doi:10.1002/jts.22888.
21. Hertenstein E, Trinca E, Wunderlin M, et al. Cognitive behavioral therapy for insomnia in patients with mental disorders and comorbid insomnia: A systematic review and meta-analysis. *Sleep Med Rev*. 2022;62.
22. Heywood SE, Connaughton J, Kinsella R, Black S, Bicchi N, Setchell J. Physical therapy and mental health: a scoping review. *Phys Ther*. 2022;102(11).
23. Hoogsteder LM, Thije L ten, Schippers EE, Stams GJJ. A meta-analysis of the effectiveness of EMDR and TF-CBT in reducing trauma symptoms and externalizing behavior problems in adolescents. *Int J Offender Ther Comp Criminol*. 2022;66(6-7):735-757.
24. Hoppen TH, Meiser-Stedman R, Jensen TK, Birkeland MS, Morina N. Efficacy of psychological interventions for post-traumatic stress disorder in children and adolescents exposed to single versus multiple traumas: meta-analysis of randomised controlled trials. *Br J Psychiatry*. 2023;222(5):196-203.
25. Huang T, Li H, Tan S, et al. The efficacy and acceptability of exposure therapy for the treatment of post-traumatic stress disorder in children and adolescents: a systematic review and meta-analysis. *BMC Psychiatry*. 2022;22(1):1-12.
26. Hudays A, Gallagher R, Hazazi A, Arishi A, Bahari G. Eye Movement Desensitization and Reprocessing versus Cognitive Behavior Therapy for Treating Post-Traumatic Stress Disorder: A Systematic Review and Meta-Analysis. *Int J Environ Res Public Health*. 2022;19(24).
27. Hutchinson R, King N, Majumder P. How effective is group intervention in the treatment for unaccompanied and accompanied refugee minors with mental health difficulties: A systematic review. *Int J Soc Psychiatry*. 2022;68(3):484-499.
28. Kalisch LA, Lawrence KA, Baud J, Spencer-Smith M, Ure A. Therapeutic supports for neurodiverse children who have experienced interpersonal trauma: A scoping review. *Rev J Autism Dev Disord*. 2023:1-23. doi:10.1007/s40489-023-00363-9.
29. Kip A, Iseke LN, Papola D, Gastaldon C, Barbui C, Morina N. Efficacy of psychological interventions for PTSD in distinct populations-An evidence map of meta-analyses using the umbrella review methodology. *Clin Psychol Rev*. 2022;100.
30. Kularatna S, Hettiarachchi R, Senanayake S, Murphy C, Donovan C, March S. Cost-effectiveness analysis of paediatric mental health interventions: a systematic review of model-based economic evaluations. *BMC Health Serv Res*. 2022;22(1):1-17.
31. Le Roux IH, Cobham VE. Psychological Interventions for Children Experiencing PTSD After Exposure to a Natural Disaster: A Scoping Review. *Clin Child Fam Psychol Rev*. 2022;(25):249-282. doi:10.1007/s10567-021-00373-1.
32. Lee J, Dhauna J, Silvers JA, Houston MH, Barnert ES. Therapeutic dance for the healing of sexual trauma: A systematic review. *Trauma Violence Abuse*. 2023;24(4):2143-2164.
33. Lobanov-Rostovsky S, Kiss L. The mental health and well-being of internally displaced female Yazidis in the Kurdistan Region of Iraq: a realist review of psychosocial interventions and the impact of COVID-19. *GLOB MENT HEALTH*. 2022;9:1-13.
34. Lotzin A, Franc de Pommereau A, Laskowsky I. Promoting recovery from disasters, pandemics, and trauma: a systematic review of brief psychological interventions to reduce distress in adults, children, and adolescents. *Int J Environ Res Public Health*. 2023;20(7). doi:10.3390/ijerph20075339.
35. Macgowan MJ, Naseh M, Rafieifar M. Eye Movement Desensitization and Reprocessing to Reduce Post-Traumatic Stress Disorder and Related Symptoms among Forcibly Displaced People: A Systematic Review and Meta-Analysis. *Res Soc Work Pract*. 2022;32(8):863-877.
36. Mavranezouli I, Megnin‐Viggars O, Daly C, et al. Research Review: Psychological and psychosocial treatments for children and young people with post‐traumatic stress disorder: a network meta‐analysis. *J Child Psychol Psychiatry*. 2020;61(1):18-29.
37. Mazzeo G, Bendixen R. Community-Based Interventions for Childhood Trauma: A Scoping Review. *OTJR*. 2023;43(1):14-23. doi:10.1177/15394492221091718.
38. Mendlowicz MV, Gekker M, Xavier Gomes de Araújo, Alexandre, et al. The top-100 cited articles on post-traumatic stress disorder: a historical bibliometric analysis. *Psychol Health Med*. 2022:1-20.
39. Mitchell S, Mitchell R, Shannon C, Dorahy M, Hanna D. Effects of baseline psychological symptom severity on dropout from trauma-focused cognitive behavior therapy for posttraumatic stress disorder: A meta-analysis. *Traumatology*. 2023;29(2):112-124.
40. Morison L, Simonds L, Stewart S-JF. Effectiveness of creative arts-based interventions for treating children and adolescents exposed to traumatic events: A systematic review of the quantitative evidence and meta-analysis. *Arts Health*. 2022;14(3):237-262.
41. Oystrick V, Coholic D, Schinke R. A Scoping Review of Mindfulness-Based and Arts-Based Parenting Interventions for Adolescent Mothers. *Child Adolesc Social Work J*. 2023:1-23. doi:10.1007/s10560-023-00923-2.
42. Ozamiz-Etxebarria N, Legorburu Fernandez I, Idoiaga-Mondragon N, Olaya B, Cornelius-White JHD, Santabárbara J. Post-Traumatic Stress in Children and Adolescents during the COVID-19 Pandemic: A Meta-Analysis and Intervention Approaches to Ensure Mental Health and Well-Being. *Sustainability*. 2023;15(6). doi:10.3390/su15065272.
43. Paggiaro AO, Paggiaro PBS, Fernandes RAQ, Freitas NO, Carvalho VF, Gemperli R. Posttraumatic stress disorder in burn patient: a systematic review. *J Plast Reconstr Aesthet Surg*. 2022;75(5):1586-1595.
44. Peters W, Rice S, Alvarez‐Jimenez M, et al. Relative efficacy of psychological interventions following interpersonal trauma on anxiety, depression, substance use, and PTSD symptoms in young people: A meta‐analysis. *Early Interv Psychiatry*. 2022;16(11):1175-1184.
45. Ramírez-Guarín V, Codina N, Pestana JV. A systematic review of psychosocial interventions for child soldiers: types, length and main findings. *J Soc Work Pract*. 2023;37(1):79-95. doi:10.1080/02650533.2022.2031934.
46. Roche L, McLay L, Sigafoos J, Whitcombe‐Dobbs S. A review of behavioral treatments for sleep disturbances in civilians who have experienced trauma. *Behav Interv*. 2022;37(3):835-863.
47. Rossouw J, Sharp T, Halligan S, Seedat S. Psychotherapeutic interventions for childhood posttraumatic stress disorder: an update. *Curr Opin Psychiatry*. 2022;35(6):417-424.
48. Samarah EMS. Narrative exposure therapy to address PTSD symptomology with refugee and migrant children and youth: A review. *Traumatology*:in print.
49. Semmlinger V, Ehring T. Predicting and preventing dropout in research, assessment and treatment with refugees. *Clin Psychol Psychother*. 2022;29(3):767-782.
50. Soltan F, Cristofalo D, Marshall D, et al. Community‐based interventions for improving mental health in refugee children and adolescents in high‐income countries. *Cochrane Database Syst Rev*. 2022;(5):CD013657.
51. Somers K, Spruit A, Stams GJ, Vandevelde S, Lindauer R, Assink M. Identifying effective moderators of cognitive behavioural trauma treatment with caregiver involvement for youth with PTSD: A meta-analysis. *Eur Child Adolesc Psychiatry*:in print.
52. Stewart TM, Fry D, Wilson J, et al. Adolescent Mental Health Priorities During the Covid-19 Pandemic. *Sch Ment Health*. 2023;15(1):247-259.
53. Sunderland N, Stevens F, Knudsen K, Cooper R, Wobcke M. Trauma aware and anti-oppressive arts-health and community arts practice: Guiding principles for facilitating healing, health and wellbeing. *Trauma Violence Abuse*. 2023;24(4):2429-2447.
54. Szota K, Schulte KL, Christiansen H. Interventions involving caregivers for children and adolescents following traumatic events: A systematic review and meta-analysis. *Clin Child Fam Psychol Rev*. 2023;26(1):17-32. doi:10.1007/s10567-022-00415-2.
55. Thielemann JF, Kasparik B, König J, Unterhitzenberger J, Rosner R. A systematic review and meta-analysis of trauma-focused cognitive behavioral therapy for children and adolescents. *Child Abuse Negl*. 2022;134:105899. doi:10.1016/j.chiabu.2022.105899.
56. Thomas FC, Puente‐Duran S, Mutschler C, Monson CM. Trauma‐focused cognitive behavioral therapy for children and youth in low and middle‐income countries: A systematic review. *Child Adolesc Ment Health*. 2022;27(2):146-160.
57. Trubey R, Evans R, McDonald S, et al. Effectiveness of Mental Health and Wellbeing Interventions for Children and Young People in Foster, Kinship, and Residential Care: Systematic Review and Meta-Analysis. *Trauma Violence Abuse*:in print.
58. van der Hoeven ML, Assink M, Stams G-JJM, Daams JG, Lindauer RJL, Hein IM. Victims of child abuse dropping out of trauma-focused treatment: a meta-analysis of risk factors. *J Child Adolesc Trauma*. 2023;16(2):269-283.
59. Venturo-Conerly KE, Eisenman D, Wasil AR, Singla DR, Weisz JR. Meta-analysis: the effectiveness of youth psychotherapy interventions in low-and middle-income countries. *J Am Acad Child Adolesc Psychiatry*. 2023;62(8):859-873.
60. Vieira S, Liang X, Guiomar R, Mechelli A. Can we predict who will benefit from cognitive-behavioural therapy? A systematic review and meta-analysis of machine learning studies. *Clin Psychol Rev*. 2022;97:102193.
61. Villarreal-Zegarra D, Alarcon-Ruiz CA, Melendez-Torres GJ, et al. Development of a framework for the implementation of synchronous digital mental health: realist synthesis of systematic reviews. *JMIR Ment Health*. 2022;9(3):e34760.
62. Wamser‐Nanney R, Walker HE. Attrition from pediatric trauma‐focused cognitive behavioral therapy: A meta‐analysis. *J Trauma Stress*. 2023;36(1):17-30. doi:10.1002/jts.22890.
63. Wang W, Chen K, Zhang H. Effectiveness of Trauma-Focused Cognitive Behavioral Therapy Among Maltreated Children: A Meta-Analysis. *Res Soc Work Pract*. 2023;33(8):913-928. doi:10.1177/104973152211472.
64. Wichmann MLY, Pawils S, Richters J, Metzner F. School-based interventions for child and adolescent victims of interpersonal violence. *Trauma Violence Abuse*. 2023;24(3).
65. Willmot RA, Sharp RA, Kassim AA, Parkinson JA. A scoping review of community-based mental health intervention for children and adolescents in South Asia. *Glob Ment Health*. 2023;10(1):e1. doi:10.1017/gmh.2022.49.
66. Xian-Yu C-Y, Deng N-J, Zhang J, et al. Cognitive behavioral therapy for children and adolescents with post-traumatic stress disorder: meta-analysis. *J Affect Disord*. 2022;308:502-511.
67. Xie S, Cheng Q, Tan S, et al. The efficacy and acceptability of group trauma-focused cognitive behavior therapy for the treatment of post-traumatic stress disorder in children and adolescents: A systematic review and meta-analysis. *General Hospital Psychiatry*. 2024;86:127-134.
68. Xiang Y, Cipriani A, Teng T, et al. Comparative efficacy and acceptability of psychotherapies for post-traumatic stress disorder in children and adolescents: a systematic review and network meta-analysis. *BMJ Ment Health*. 2021;24(4):153-160.
69. Xiong T, Milios A, McGrath PJ, Kaltenbach E. The influence of social support on posttraumatic stress symptoms among children and adolescents: a scoping review and meta-analysis. *Eur J Psychotraumatol*. 2022;13(1):2011601.
70. Yohannan J, Carlson JS, Volker MA. Cognitive behavioral treatments for children and adolescents exposed to traumatic events: A meta‐analysis examining variables moderating treatment outcomes. *J Trauma Stress*. 2022;35(2):706-717.
71. Yosep, I., Mardhiyah, A., Ramdhanie, G. G., Sari, C. W. M., Hendrawati, H., & Hikmat, R. Cognitive Behavior Therapy by Nurses in Reducing Symptoms of Post-Traumatic Stress Disorder on Children as Victims of Violence: A Scoping Review. *Healthcare*. 2023;11(3):e407. https://www.mdpi.com/2227-9032/11/3/407. Published January 31, 2023.
72. Young G. Psychotherapeutic change mechanisms and causal psychotherapy: Applications to child abuse and trauma. *J Child Adolesc Trauma*. 2022;15(3):911-923.
73. Zadeh R, Jogia J. The Use of Art Therapy in Alleviating Mental Health Symptoms in Refugees: A Literature Review. *Int J Ment Health Promot*. 2023;25(3):309-326. doi:10.32604/ijmhp.2023.022491.
74. Zisopoulou T, Varvogli L. Stress Management Methods in Children and Adolescents: Past, Present, and Future. *Horm Res Paediatr*. 2023;96(1):97-107.

**Appendix C: Quality criteria for risk of bias assessment**

Quality criteria based on Cuijpers et al. (2010), sum score ranges from 0 to 8, high-quality trials fulfilled at least six of these eight criteria.

| 1. All participants met diagnostic criteria for PTSD at baseline as assessed via a diagnostic interview based on any iteration of the DSM or ICD | 1. Positive   0. Negative / insufficient information |
| --- | --- |
| 2. Use of treatment manual   *(i.e., published, or specifically designed for the study;* ***all*** *psychological interventions of the RCT/included in the analyses were manual-based*  *🡪 insufficient: manual mentioned but without a reference/specification)* | 1. Positive   0. Negative / insufficient information |
| 3. Therapists were trained in applying the manual   *(i.e., specifically for the study or general training for respective manual)* | 1. Positive   0. Negative / insufficient information |
| 4. Treatment integrity was checked formally   *(i.e., by regular supervision and/or recordings and/or systematic screenings of protocol adherence with a standardized instrument)* | 1. Positive   0. Negative / insufficient information |
| 5. Data analyzed with intent-to-treat analysis   *(i.e., all persons who were randomized to the conditions initially were included in analyses; insufficient: if authors stated that both ITT and completer analyses were performed but only reported on completer results)* | 1. Positive   0. Negative / insufficient information |
| 6. Study had an adequate level of statistical power to find effects and included *N* ≥ 50 participants in the comparison groups (i.e., *n1+n2*)   *(note: may differ across assessment timepoints due to attrition in completer analyses)* | 1. Positive   0. Negative / insufficient information |
| 7. Randomization by independent (3^rd^) party   *(e.g., independent person or computerized with valid randomization technique)* | 1. Positive   0. Negative / insufficient information |
| 8. Blind assessors for outcomes   *(i.e., PTSD outcomes were assessed either in a diagnostic interviewer with blinded assessors or via a self-report measure; negative score: non-blinded interviewers)* | 1. Positive   0. Negative / insufficient information |

**Appendix D: Quality coding of included trials**

|  | Q1 | Q2 | Q3 | Q4 | Q5 | Q6 | Q7 | Q8 | Quality sum score  (possible range = 0-8) |
| --- | --- | --- | --- | --- | --- | --- | --- | --- | --- |
| Reference | 100% PTSD rate at baseline | manual-based treatment | therapists trained to apply manual | formal integrity check | usable intent-to-treat (ITT) data reported | *N* > 50, high power to detect effects | independent randomization | blinded outcome assessments |  |
| Ahmad et al. (2007) | 1 | 1 | 0 | 0 | 1 | 0 | 0 | 1 | 4 |
| **Ahmadi et al. (2022) Post** | **0** | **1** | **1** | **1** | **0** | **1** | **1** | **1** | **6** |
| Ahmadi et al. (2022) FU 1 | 0 | 1 | 1 | 1 | 0 | 0 | 1 | 1 | 5 |
| Ahrens & Rexford (2002) | 0 | 1 | 1 | 0 | 0 | 0 | 0 | 1 | 3 |
| Al-Hadethe et al. (2015) | 0 | 0 | 0 | 0 | 0 | 0 | 1 | 1 | 2 |
| Auslander et al. (2017) | 0 | 1 | 1 | 1 | 0 | 0 | 1 | 1 | 5 |
| **Auslander et al. (2020)** | **0** | **1** | **1** | **1** | **1** | **1** | **1** | **0** | **6** |
| **Banoglu & Korkmazlar (2021)** | **1** | **1** | **1** | **1** | **0** | **1** | **0** | **1** | **6** |
| **Barron et al. (2016)** | **0** | **1** | **1** | **1** | **1** | **1** | **1** | **1** | **7** |
| Barron et al. (2021) | 0 | 1 | 1 | 1 | 1 | 0 | 0 | 1 | 5 |
| **Bidstrup et al. (2021)** | **0** | **1** | **1** | **1** | **1** | **0** | **1** | **1** | **6** |
| **Carrion et al. (2013)** | **0** | **1** | **1** | **1** | **1** | **1** | **1** | **1** | **7** |
| **Catani et al. (2009)** | **0** | **1** | **1** | **1** | **1** | **0** | **1** | **1** | **6** |
| Celano et al. (1996) | 0 | 1 | 1 | 1 | 0 | 0 | 0 | 1 | 4 |
| Chemtob et al. (2002) | 1 | 1 | 1 | 1 | 0 | 0 | 0 | 1 | 5 |
| Chen et al. (2014) | 0 | 1 | 0 | 0 | 0 | 0 | 0 | 1 | 2 |
| Cohen et al. (2004) | 0 | 1 | 1 | 1 | 0 | 1 | 0 | 1 | 5 |
| **Cohen et al. (2005)** | **0** | **1** | **0** | **1** | **1** | **1** | **1** | **1** | **6** |
| **Cohen et al. (2011)** | **0** | **1** | **1** | **1** | **1** | **1** | **1** | **1** | **7** |
| Danielson et al. (2012) | 0 | 0 | 1 | 1 | 1 | 0 | 1 | 1 | 5 |
| Dawson et al. (2018) | 0 | 0 | 1 | 0 | 1 | 1 | 1 | 1 | 5 |
| **de Roos et al. (2011)** | **0** | **1** | **1** | **1** | **1** | **0** | **1** | **1** | **6** |
| **de Roos et al. (2017)** | **0** | **1** | **1** | **1** | **1** | **1** | **1** | **1** | **7** |
| Deblinger et al. (1996) | 0 | 1 | 1 | 1 | 0 | 0 | 0 | 0 | 3 |
| **Diehle et al. (2015)** | **0** | **1** | **1** | **1** | **1** | **0** | **1** | **1** | **6** |
| **Dorsey et al. (2020) Kenya urban** | **0** | **1** | **1** | **1** | **1** | **1** | **1** | **1** | **7** |
| **Dorsey et al. (2020) Kenya rural** | **0** | **1** | **1** | **1** | **1** | **1** | **1** | **1** | **7** |
| **Dorsey et al. (2020) Tanzania urban Post** | **0** | **1** | **1** | **1** | **1** | **1** | **1** | **1** | **7** |
| **Dorsey et al. (2020) Tanzania urban FU 2** | **0** | **1** | **1** | **1** | **1** | **0** | **1** | **1** | **6** |
| **Dorsey et al. (2020) Tanzania rural** | **0** | **1** | **1** | **1** | **1** | **0** | **1** | **1** | **6** |
| **El-Khani et al. (2021)** | **0** | **1** | **1** | **1** | **1** | **1** | **1** | **1** | **7** |
| **Ertl et al. (2011)** | **1** | **1** | **1** | **1** | **0** | **1** | **0** | **1** | **6** |
| Espil et al. (2022) | 0 | 0 | 1 | 1 | 0 | 0 | 0 | 0 | 2 |
| **Foa et al. (2013)** | **0** | **1** | **1** | **1** | **1** | **1** | **1** | **1** | **7** |
| **Ford et al. (2012)** | **0** | **1** | **1** | **1** | **1** | **1** | **1** | **0** | **6** |
| **Gilboa-Schechtman et al. (2010)** | **1** | **1** | **1** | **1** | **1** | **0** | **1** | **1** | **7** |
| **Goldbeck et al. (2016)** | **0** | **1** | **1** | **1** | **1** | **1** | **1** | **1** | **7** |
| **Gordon et al. (2008)** | **0** | **1** | **1** | **0** | **1** | **1** | **1** | **1** | **6** |
| **Hitchcock et al. (2021) study 2** | **1** | **1** | **1** | **1** | **1** | **0** | **1** | **1** | **7** |
| **Jensen et al. (2014)** | **0** | **1** | **1** | **1** | **1** | **1** | **1** | **1** | **7** |
| **Kameoka et al. (2020)** | **0** | **1** | **1** | **1** | **1** | **0** | **1** | **1** | **6** |
| **Kaminer et al. (2023)** | **0** | **1** | **1** | **1** | **1** | **1** | **1** | **1** | **7** |
| Kemp et al. (2010) | 0 | 1 | 1 | 1 | 0 | 0 | 0 | 0 | 3 |
| King et al. (2000) | 0 | 1 | 1 | 1 | 1 | 0 | 0 | 0 | 4 |
| **Langley et al. (2015)** | **0** | **1** | **1** | **1** | **0** | **1** | **1** | **1** | **6** |
| Lesmana et al. (2009) | 0 | 0 | 0 | 0 | 1 | 1 | 0 | 0 | 2 |
| **Li et al. (2023)** | **0** | **1** | **1** | **1** | **1** | **1** | **1** | **1** | **7** |
| **Li et al. (2024)** | **0** | **1** | **1** | **1** | **1** | **1** | **1** | **1** | **7** |
| McMullen et al. (2013) | 0 | 1 | 1 | 1 | 0 | 0 | 1 | 1 | 5 |
| **Meentken et al. (2020)** | **0** | **1** | **0** | **1** | **1** | **1** | **1** | **1** | **6** |
| Meiser-Stedman et al. (2017) | 1 | 1 | 1 | 1 | 0 | 0 | 0 | 1 | 5 |
| Molero et al. (2019) | 0 | 1 | 1 | 0 | 0 | 1 | 1 | 1 | 5 |
| **Murray et al. (2015)** | **0** | **1** | **1** | **1** | **1** | **1** | **1** | **1** | **7** |
| **O'Callaghan et al. (2013)** | **0** | **1** | **1** | **1** | **1** | **1** | **1** | **1** | **7** |
| **O'Callaghan et al. (2015)** | **0** | **1** | **1** | **1** | **1** | **1** | **1** | **1** | **7** |
| **Osorio et al. (2018)** | **0** | **1** | **1** | **1** | **1** | **0** | **1** | **1** | **6** |
| Peltonen et al. (2019) | 0 | 1 | 1 | 1 | 0 | 0 | 1 | 0 | 4 |
| Pityaratstian et al. (2015) | 1 | 1 | 1 | 0 | 0 | 0 | 0 | 1 | 4 |
| Roque-Lopez et al. (2021) | 0 | 1 | 1 | 0 | 0 | 0 | 1 | 0 | 3 |
| **Rosner et al. (2019)** | **1** | **1** | **1** | **1** | **1** | **1** | **1** | **1** | **8** |
| **Rossouw et al. (2020)** | **1** | **1** | **1** | **1** | **1** | **1** | **1** | **1** | **8** |
| Ruf et al. (2010) | 1 | 1 | 1 | 0 | 1 | 0 | 0 | 1 | 5 |
| Santiago et al. (2018) | 0 | 1 | 1 | 0 | 1 | 1 | 0 | 1 | 5 |
| **Schauer (2008)** | **1** | **1** | **1** | **1** | **1** | **0** | **1** | **1** | **7** |
| Scheeringa et al. (2011) | 0 | 1 | 0 | 1 | 0 | 0 | 1 | 0 | 3 |
| Schottelkorb et al. (2012) | 0 | 1 | 1 | 1 | 0 | 0 | 1 | 0 | 4 |
| Shechtman & Mor (2010) | 0 | 1 | 1 | 1 | 0 | 1 | 0 | 1 | 5 |
| **Shein-Szydlo et al. (2016)** | **1** | **1** | **1** | **1** | **0** | **1** | **1** | **1** | **7** |
| Sloan et al. (2011) | 1 | 1 | 1 | 1 | 0 | 0 | 0 | 1 | 5 |
| **Smith et al. (2007)** | **1** | **1** | **1** | **1** | **1** | **0** | **1** | **1** | **7** |
| **Stein et al. (2003)** | **0** | **1** | **1** | **1** | **0** | **1** | **1** | **1** | **6** |
| Trowell et al. (2002) | 0 | 1 | 1 | 1 | 0 | 1 | 0 | 0 | 4 |

**Bold** font indicates that the given trial was categorized as high-quality trial (i.e., fulfilling at least six of the eight quality criteria).

**Appendix E: Categorization of interventions and control conditions**

| Overarching category | Families of psychological interventions (top) and control group categories (bottom) | Specific interventions/manuals for each family of interventions (top) and specific control conditions for control condition categories |
| --- | --- | --- |
| Psychological interventions | Trauma-focused cognitive behavior therapy interventions (TF-CBTs) | Cognitive behavioral intervention for trauma in schools (CBITS), cognitive behavior therapy for trauma in street children (CBT-TSD), trauma-based cognitive therapy protocol for young children (CBT-3M), cognitive behavior writing therapy (CBWT), cognitive processing therapy (CPT), cognitive therapy for PTSD (CT-PTSD), developmentally-adapted cognitive processing therapy (D-CPT), narrative exposure therapy for children (KIDNET), modified written exposure therapy (m-WET), narrative exposure therapy (NET), exposure therapy, power up children's psychological immunity (PCPI), prolonged exposure (PE), prolonged exposure for adolescents (PE-A), recovering from abuse program (RAP), teaching recovery techniques (TRT), teaching recovery techniques (TRT+) manual adapted to disasters, teaching recovery techniques plus parenting (TRT+P), trauma-focused cognitive behavior therapy (TF-CBT), written exposure therapy (WET). |
|  | Eye movement desensitization and reprocessing (EMDR) | Various adapted manuals (e.g., EMDR, EMDR-group protocol for children or EMDR-Integrative group treatment protocol for ongoing traumatic stress) but all labelled EMDR. |
|  | Other trauma-focused interventions (o-TF interventions) | Psychodynamic psychotherapy (PDP), expressive supportive group intervention, spiritual hypnosis-assisted treatment. |
|  | Non-trauma-focused interventions (non-TF interventions) | Child-centered therapy, child-centered play therapy, non-directive supportive therapy, problem solving therapy, emotional freedom techniques (EFT),  family-oriented support (FAMOS), meditation. |
|  | Multi-disciplinary treatments (MDTs) | Bounce back, cue-centered therapy, risk reduction through family therapy (RRFT), trauma affect regulation: Guide for education and therapy (TARGET), eclectic psychosocial intervention, intensive multimodal group  program, mind body skills group. |
| Control conditions | Active control conditions | Care-as-usual (CAU), neutral writing, psychoeducation, supportive counselling, treatment-as-usual (TAU), usual care. |
|  | Passive control conditions | Waitlist control conditions (i.e., assessments only). |

**Appendix F: Trial characteristics of included trials**

| **Publication,**  **Categorization of arms**  **(& number of sessions)** | **n** | **Intervention**  **(format of delivery, parent involvement)** | **Country of conduct** | **Age range (Mean)** | **% of total sample identifying as female** | **% of total sample fulfilling full PTSD diagnosis at baseline (measure)** | **Outcome measure (PTSD)** | **ITT data vs. completer data reported** | **Trauma type(s) experienced** | **Quality sum score (out of 8)** |
| --- | --- | --- | --- | --- | --- | --- | --- | --- | --- | --- |
| Ahmad et al. (2007)  EMDR (8 sessions, 45 min.)  PCC/WL | 17  16 | EMDR  (individual) | Sweden | 6–16  (10.0) | 60.61 | 100 (DICA) | PTSS-C | ITT | Types varied between participants | 4 |
| Ahmadi et al. (2022) TF-CBT (5 sessions)  PCC/WL | 34  38 | m-WET  (group) | Afghanistan | 12-18  (16.05) | 100 | 100 (CRIES) | CRIES | Compl. | School terrorist attack | 6 |
| Ahrens & Rexford (2002)  TF-CBT (8 sessions, 60 min.)  PCC/WL | 19  19 | CPT  (individual) | USA | 15–18  (16.4) | 0 | 100 (PSS-SR) | PSS-SR | Compl. | Physical violence | 3 |
| Al-Hadethe et al. (2015)  TF-CBT  Other  PCC/WL | 19  20  20 | NET, EFT (individual in both conditions) | Iraq | 16-19 | 0 | 100 (SPTSS) | SPTSS | Compl. | War | 2 |
| Auslander et al. (2017)  TF-CBT (10 sessions, 90 min.)  ACC/TAU (n.r.) | 15  10 | CBITS  (group and individual mixed, parent involvement) | USA | 12-18  (14.7) | 100 | 67 (CPSS) | CPSS | Compl. | Types varied between participants | 5 |
| Auslander et al. (2020)  TF-CBT (14 sessions, 90 min)  ACC/UC (n.r.) | 115  108 | CBITS  (group and individual mixed, parent involvement) | USA | 12-19  (14.89) | 100 | 54 (CPSS) | CPSS | ITT | Types varied between participants | 6 |
| Banoglu & Korkmazlar (2021) EMDR (3-4 sessions, 90-120 min.)  PCC/WL | 42  19 | EMDR-GP/C  (group) | Turkey  (refugees) | 6-15 | 41 | 59.31  (CPTS-RI) | CPTS-RI | Compl. | War | 6 |
| Barron et al. (2016)  TF-CBT (5 sessions, 60 min.)  PCC/WL | 79  75 | TRT  (group) | Palestine | 11-18  (13.5) | 59.71 | n.r. | CRIES | ITT | War | 7 |
| Barron et al. (2021)  TF-CBT (5 sessions, 90 min.) PCC/WL | 14  16 | TRT  (group) | Brazil | 8-13  (10.1) | 46.67 | 96.67 (CRIES) | CRIES | ITT | Drug violence | 3 |
| Bidstrup et al. (2021)  Non-TF-PI (7 sessions, 60-90 min.)  ACC/UC (n.r., 60 min.) | 51  58 | FAMOS  (group and individual mixed, parent involvement) | Denmark | 2-5  (3.45) | 59 | 91.5  (PEDS) | PEDS | ITT | Cancer | 6 |
| Carrion et al. (2013)  MDT (15 sessions, 50 min.)  PCC/WL | 38  27 | Cue-centered treatment (individual, parent involvement) | USA | 8–17  (11.56) | 40.00 | n.r. | UCLA PTSD-RI | ITT | Physical violence | 7 |
| Catani et al. (2009)  TF-CBT (6 sessions, 60-90 min.)  Meditation (6 sessions, 60-90 min.) | 16  15 | KIDNET  (individual) | Sri Lanka | 8–14  (11.95) | 45.16 | 100 (UPID) | UPID | ITT | Types varied between participants | 6 |
| Celano et al. (1996)  TF-CBT (8 sessions, 60 min.)  ACC/TAU (8 sessions, 60 min.) | 15  17 | Recovering from abuse program  (individual, parent involvement) | USA | 8–13  (10.5) | 100 | n.r. | CITES-R | Compl. | Sexual assault | 4 |
| Chemtob et al. (2002)  EMDR (3 sessions, n.r.)  PCC/WL | 17  15 | EMDR  (individual) | USA | 6–12  (8.4) | 68.75 | 100 (CRI) | CRI | Compl. | Disaster | 5 |
| Chen et al. (2014)  TF-CBT (6 sessions, 60 min.)  ACC/Support  PCC/WL | 10  10  12 | TRT+  (group) | China | n.a.  (14.50) | 68.00 | 100 (CRIES) | CRIES | Compl. | Parental death and earthquake | 2 |
| Cohen et al. (2004)  TF-CBT (12 sessions, 45 min.)  Non-TF-PI (12 sessions, 45 min.) | 89  91 | TF-CBT,  child-centered  therapy (individual, parent involvement in  both conditions) | USA | 8–14  (10.7) | 78.82 | 89 (K-SADS) | K-SADS | Compl. | Sexual assault | 5 |
| Cohen, et al. (2005)  TF-CBT (12 sessions, 45 min.)  Non-TF-PI (12 sessions, 45 min.) | 41  41 | TF-CBT, non-directive supportive therapy  (individual, parent involvement in both conditions) | USA | 8–15  (11.4) | 68.29 | n.r. | TSCC | ITT | Sexual assault | 6 |
| Cohen et al. (2011)  TF-CBT (8 sessions, 45 min.)  Non-TF-PI (8 sessions, 45 min.) | 64  60 | TF-CBT,  child-centered  therapy (individual, parent involvement in both conditions) | USA | 7–14  (9.6) | 50.81 | n.r. | K-SADS | ITT | Types varied between participants | 7 |
| Danielson et al. (2012)  MDT (23 sessions, 60-90 min.)  ACC/TAU (13 sessions, n.r.) | 15  15 | RRFT  (individual, parent involvement) | USA | 13–17  (14.8) | 88.00 | n.r. | UPID | ITT | Sexual assault | 5 |
| Dawson et al. (2018)  TF-CBT (6 sessions, 60 min.)  Non-TF-PI (6 sessions, 60 min.) | 32  32 | TF-CBT, PST  (individual &  parent involvement  in both conditions) | Indonesia | 7-14  (10.7) | 48.44 | n.r. | UCLA PTSD-RI | ITT | War and tsunami | 5 |
| de Roos et al. (2011)  TF-CBT (4 sessions, 60 min.)  EMDR (4 sessions, 60 min.) | 26  26 | TF-CBT,  EMDR (individual, parent involvement in both conditions) | NL | 4–18  (10.1) | 44.23 | n.r. | UCLA PTSD-RI | ITT | Explosion | 6 |
| de Roos et al. (2017)  EMDR (6 sessions, 45 min.)  TF-CBT (6 sessions, 45 min.)  PCC/WL | 43  42  18 | EMDR,  CBWT  (individual) | NL | 8-18  (13.06) | 57.28 | 61.2 (CRTI) | CRTI | ITT | Types varied between participants | 7 |
| Deblinger et al. (1996)  TF-CBT-c (12 sessions, 45 min.)  TF-CBT-c&p (12 sessions 90 min.)  ACC/TAU (n.r.) | 24  22  22 | TF-CBT  (individual, child only vs. child & parent) | USA | 7–13  (9.8) | 83.00 | 71 (K-SADS) | K-SADS | Compl. | Sexual assault | 3 |
| Diehle et al. (2015)  TF-CBT (12 sessions, 60 min.)  EMDR (12 sessions, 60 min.) | 23  25 | TF-CBT,  EMDR  (individual, parent involvement in both conditions) | NL | 8–18  (12.9) | 62.50 | 50 (ADIS-P) | CAPS-CA | ITT | Types varied between participants | 6 |
| Dorsey et al. (2020)  TF-CBT (15 sessions, n.r.)  ACC/UC (n.r.) | 96  96 | TF-CBT  (individual and group mixed) | Kenya  /urban | 7-13  (10.7) | 50 | 100 (CPSS) | CPSS | ITT | Parental death and other types | 7 |
| Dorsey et al. (2020)  TF-CBT (15 sessions, n.r.)  ACC/UC (n.r.) | 64  64 | TF-CBT  (individual and group mixed) | Kenya  /rural | 7-13  (10.2) | 50 | 100 (CPSS) | CPSS | ITT | Parental death and other types | 7 |
| Dorsey et al. (2020)  TF-CBT (15 sessions, n.r.)  ACC/UC (n.r.) | 96 96 | TF-CBT  (individual and group mixed) | Tanzania /urban | 7-13 (10.9) | 50 | 100 (CPSS) | CPSS | ITT | Parental death and other types | 7 |
| Dorsey et al. (2020)  TF-CBT (15 sessions, n.r.)  ACC/UC (n.r.) | 64  64 | TF-CBT  (individual and group mixed) | Tanzania/  rural | 7-13  (10.5) | 50 | 100 (CPSS) | CPSS | ITT | Parental death and other types | 7 |
| El-Khani et al. (2021)  TF-CBT (7 sessions, 120 min.)  PCC/WL | 41  40 | TRT+P  (group, parent involvement) | Libanon (refugees) | 9-12 | 87.65 | 100 (CRIES) | CRIES | ITT | War | 7 |
| Ertl et al. (2011)  TF-CBT (8 sessions, 90-120 min.)  ACC/SC (8 sessions, 90-120 min.)  PCC/WL | 26  24  28 | KIDNET (individual) | Uganda | 12-25 (18.0) | 55.29 | 100 (CAPS-CA) | CAPS-CA | Compl. | War | 6 |
| Espil et al. (2022) TF-CBT (15-18 sessions, 50 min.)  MDT (15-18 sessions, 50 min.)  ACC/TAU (15-18 sessions, 50 min.) | 22  25  26 | TF-CBT, Cue-centered therapy  (individual) | USA | 7-17  (12.97) | 62.87 | 100 (UCLA PTSD-RI) | UCLA PTSD-RI | ITT | Types varied between participants | 2 |
| Foa et al. (2013)  TF-CBT (14 sessions, 60-90 min.)  ACC/SC (14 sessions, 60-90 min.) | 31  30 | PE  (individual) | USA | 13–18  (15.3) | 100 | 100 (CPSS-I) | CPSS-I | ITT | Sexual assault | 7 |
| Ford et al. (2012)  MDT (12 sessions, 50 min.)  ACC/TAU (12 sessions, 50 min.) | 33  26 | TARGET  (individual) | USA | 13–17  (14.7) | 100 | 63 (CAPS-CA) | CAPS-CA | ITT | Types varied between participants | 6 |
| Gilboa-Schechtman et al. (2010)  TF-CBT (12–15 sessions, 60-90 min.)  Other-TF-PI (15–18 sessions, 60-90 min.) | 19  19 | PE,  PDP  (individual) | Israel | 12–18  (14.1) | 63.16 | 100 (CPSS) | CPSS | ITT | Types varied between participants | 7 |
| Goldbeck et al. (2016)  TF-CBT (12 sessions, 90 min.)  PCC/WL | 76  83 | TF-CBT  (individual,  parent involvement) | Germany | 7-17  (13.03) | 71.70 | 75.5 (CAPS-CA) | CAPS-CA | ITT | Types varied between participants | 7 |
| Gordon et al. (2008)  MDT (12 sessions, 120 min.)  PCC/WL | 38  40 | Mind-body skills group  (group) | Kosovo | 14–18  (16.3) | 75.61 | 100 (HTQ) | HTQ | Compl. | War | 7 |
| Hitchcock et al. (2021)  TF-CBT (12 sessions, n.r.) ACC/TAU (n.r.) | 18  19 | CBT-3M  (individual, parent involvement) | UK | 3-8  (6.26) | 51.35 | 100 (DIPA, diagnosis PTSD-YC) | YCPC | ITT | Types varied between participants | 6 |
| Jensen et al. (2014)  TF-CBT (12-15 sessions, 45 min.)  ACC/TAU (n.r.) | 79  77 | TF-CBT  (individual, parent involvement) | Norway | 10–18  (15.1) | 79.49 | 66.7 (CAPS-CA) | CAPS-CA (post) CPSS (FUs) | ITT | Types varied between participants | 7 |
| Kameoka et al. (2020)  TF-CBT (12 sessions, 95 min.)  PCC/WL | 14  16 | TF-CBT  (individual, parent involvement) | Japan | 6-18  (13.9) | 73.3 | 100 (K-SADS) | K-SADS | ITT | Types varied between participants | 6 |
| Kaminer et al. (2023)  TF-CBT (8 sessions, 90 min.)  ACC/TAU (n.r., n.r.) | 37  38 | TF-CBT  (individual, parent involvement) | South Africa | 11-19 (14.92) | 72 | 81.3 (CPSS-5) | CPSS-5 | ITT | Types varied between participants | 7 |
| Kemp et al. (2010)  EMDR (4 sessions, 60 min.)  PCC/WL | 12  12 | EMDR  (individual) | Australia | 6–12  (8.9) | 44.44 | n.r. | CPTS-RI | Compl. | Motor vehicle accident | 3 |
| King et al. (2000)  TF-CBT-c (20 sessions, 50 min.)  TF-CBT-c&p (20 sessions, 50 min.)  PCC/WL | 12  12  12 | TF-CBT  (individual, child only, child & parent involvement) | Australia | 5–17  (11.5) | 69.44 | 69 (ADIS) | ADIS | ITT | Sexual assault | 4 |
| Langley et al. (2015)  MDT (10 group+2-3 individual sessions, 50-60 min. + 30-50 min.)  PCC/WL | 35  36 | Bounce Back  (individual  and group mixed,  parent involvement) | USA | 1st-5th  grade  (7.7) | 50.00 | n.r. | UPID | Compl. | Types varied between participants | 6 |
| Lesmana et al. (2009)  Other-TF-PI (1 session, 30 min.)  PCC/WL | 48  178 | SHAT  (group) | Indonesia | 6-12  (9.83) | 52.70 | 100 (n.r.) | Self-developed measure based on DSM-IV-TR criteria | ITT | Terrorist attack | 2 |
| Li et al. (2023)  TF-CBT (10 to 12 sessions, up to 50 min.)  ACC/TAU (36 sessions, 45 min.) | 118  116 | PCPI  (group and individual mixed) | China | 9-12  (10.41) | 41.45% | 82.91% (PCL-5) | UCLA PTSD-RI-5 | ITT | Types varied between participants | 7 |
| Li et al. (2024)  TF-CBT (10 sessions, up to 50 min.)  ACC/TAU (n.r., 45 min.) | 45  42 | PCPI  (group and individual mixed) | China | 9-12  (11.00) | 65.52% | 65.52% (PCL-5) | UCLA PTSD-RI-5 | ITT | Types varied between participants | 7 |
| McMullen et al. (2013)  TF-CBT (15 sessions, 45 min.)  PCC/WL | 24  24 | TF-CBT  (group and individual mixed) | DR Congo | 13–17  (15.8) | 0 | n.r. | UCLA PTSD-RI | Compl. | War | 5 |
| Meentken et al. (2020)  EMDR (3.5 sessions, 50 min.)  ACC/CAU (n.r.) | 37  37 | EMDR  (individual, parent involvement) | NL | 4-15  (9.6) | 33.78 | 100 (CAPS-CA) | CRTI | ITT | Types varied between participants | 6 |
| Meiser-Stedman et al. (2017)  TF-CBT (10 sessions, 90 min.)  PCC/WL | 14  15 | CT-PTSD  (individual) | UK | 8-17  (13.3) | 72.41 | 75.9 (CPTSDI) | CPTSDI | ITT | Types varied between participants | 5 |
| Molero et al. (2019)  EMDR (9 sessions, first session 95 min., subsequent sessions 50 min.)  PCC/WL | 30  33 | EMDR-IGTP-OTS  (group) | Spain | 13-17  (16.36) | 0 | n.r. | PCL-5 | Compl. | Types varied between participants | 5 |
| Murray et al. (2015)  TF-CBT (10-16 sessions, 60-90 min.)  ACC/TAU (n.r.) | 131  126 | TF-CBT (individual) | Zambia | 5-18  (13.6) | 50.97 | n.r. | UPID | ITT | Types varied between participants | 7 |
| O'Callaghan et al. (2013)  TF-CBT (15 sessions, 45 min.)  PCC/WL | 24  28 | TF-CBT  (group and individual mixed, parent involvement) | DR Congo | 12–17  (16.1) | 100 | 60 (UCLA  PTSD-RI) | UCLA PTSD-RI | ITT | War and sexual assault | 7 |
| O'Callaghan et al. (2015)  TF-CBT (9 sessions, 90 min)  MDT (9 sessions, 90 min.) | 26  24 | TF-CBT,  eclectic psychosocial intervention  (group, parent involvement in  both conditions) | DR Congo | 8–17  (14.8) | 42.00 | 92 (UCLA PTSD-RI) | UCLA PTSD-RI | ITT | War | 7 |
| Osorio et al. (2018)  EMDR (6 sessions, first session 100 min., subsequent sessions 50 min.)  PCC/WL | 11  12 | EMDR-IGTP-OTS  (group) | Mexiko | 13-21  (16.71) | 43.48 | n.r. | PCL-5 | ITT | Cancer | 6 |
| Peltonen & Kangaslampi (2019)  TF-CBT (7-10 sessions, 90 min.)  ACC/TAU (n.r., 45-90 min.) | 29  21 | NET (individual) | Finland | n.a.  (13.24) | 42.00 | n.r. | CRIES | ITT | Types varied between participants | 4 |
| Pityaratstian et al. (2015)  TF-CBT (3 sessions, 120 min.)  PCC/WL | 18  18 | TF-CBT  (group) | Thailand | 10-15  (12.3) | 72.22 | 100 (UCLA PTSD-RI) | UCLA PTSD-RI | ITT | Tsunami | 4 |
| Roque-Lopez et al. (2021)  MDT (seven days, 1-week intensive program)  ACC/TAU (seven days) | 19  17 | Intensive multimodal group  program  (group) | Colombia | 13-16  (14.05) | 100 | n.r. | SPRINT | Compl. | Types varied between participants | 3 |
| Rosner et al. (2019)  TF-CBT (30 sessions, 50 min.)  PCC/WL | 44  44 | D-CPT (individual) | Germany | 14-21  (18.1) | 85.23 | 100 (CAPS-CA) | CAPS-CA | ITT | Sexual assault and physical violence | 8 |
| Rossouw et al. (2020)  TF-CBT (7-14 sessions, 60 min.)  ACC/SC (7-14 sessions, 60 min.) | 31  32 | PE  (individual) | South Africa | 13–18  (15.35) | 87.30 | 100 (MINI-KID) | CPSS-I | ITT | Types varied between participants | 8 |
| Ruf et al. (2010)  TF-CBT (8 sessions, 90-120 min.)  PCC/WL | 13  13 | KIDNET  (individual) | Germany  (refugees) | 7–16  (11.5) | 46.15 | 100 (UPID) | UPID | ITT | War | 5 |
| Santiago et al. (2018)  MDT (10 group +2 individual sessions, 50-60 min + 30-50min.)  PCC/WL | 25  27 | Bounce Back (group and individual mixed, parent involvement) | USA | 1st-4th grade (7.76) | 36.54 | n.r. | UCLA PTSD-RI | ITT | Types varied between participants | 5 |
| Schauer (2008)  TF-CBT (6 sessions, 60-90 min.)  Non-TF-PI (6 sessions, 60-90 min.) | 25  22 | KIDNET,  Meditation  (individual) | Sri Lanka | 11-15  (13.1) | 61.70 | 100 (CAPS-CA) | CAPS-CA | ITT | War | 7 |
| Scheeringa et al. (2011)  TF-CBT (12 sessions, 45 min.)  PCC/WL | 20  11 | CBT (individual, parent involvement) | USA | 3–6  (5.3) | 33.80 | 72 (PAPA) | PAPA | Compl. | Types varied between participants | 3 |
| Schottelkorb et al. (2012)  TF-CBT (12-20 sessions, 90 min.)  Non-TF-PI (24 sessions, 30 min.) | 12  14 | TF-CBT,  CCPT  (individual,  parent involvement in both conditions) | USA  (refugees) | 6-13  (9.1) | 45.20 | 58 (UCLA) | UPID | Compl. | War | 5 |
| Shechtman & Mor (2010)  Other-TF-PI (10 sessions, n.r.)  PCC/WL | 84  52 | ESGI  (group) | Israel | 9-14 | 68.00 | n.r. | CPTS-RI | Compl. | War or parental death | 4 |
| Shein-Szydlo et al. (2016)  TF-CBT (12 sessions, 60 min.)  PCC/WL | 50  49 | CBT-TSC  (individual) | Mexico | 12-19  (14.9) | 63.64 | 100 (DISC) | CPSS | ITT | Types varied between participants | 7 |
| Sloan et al. (2011)  TF-CBT (3 sessions, 20 min.)  ACC/Neutral Writing (3 sessions, 20 min.) | 21  21 | Written Exposure Therapy (individual) | USA | n.r.  (18.90) | n.r. | 100 (PSS-I) | PSS-I | Compl. | Types varied between participants | 5 |
| Smith et al. (2007)  TF-CBT (10 sessions, n.r.)  PCC/WL | 12  12 | TF-CBT  (individual, parent involvement) | UK | 8–18  (13.8) | 39.47 | 100 (CAPS-CA) | CAPS-CA | ITT | Types varied between participants | 7 |
| Stein et al. (2003)  TF-CBT (10 sessions, 60 min.)  PCC/WL | 54  63 | TF-CBT  (group) | USA | 6th grade (11.0) | 56.35 | 100 (CPSS) | CPSS | Compl. | Types varied between participants | 6 |
| Trowell et al. (2002)  Other-TF-PI (30 sessions, 50 min.)  ACC/Psychoeducation (18 group sessions, n.r.) | 28  28 | PDP  (individual) | UK | 6–14  (10.0) | 100 | 73 (DSM-IV) | K-SADS | Compl. | Sexual assault | 4 |

*Note.* ADIS = Anxiety Disorders Interview Schedule; CAPS-CA = Clinician Administered PTSD Scale for Children and Adolescents; CAU = care-as-usual; CBT = cognitive behaviour therapy; CBT-TSC = CBT for trauma in street children; trauma-based cognitive therapy protocol for young children (CBT-3M); CCPT = Child Centered Play Therapy; CCT = Child Centered Therapy; CITES-R = Children's Impact of Traumatic Events Scales-Revised; Compl.= completer data; CPSS = Child PTSD Symptom Scale; CPSS-I = CPSS interview version; CPT = Cognitive Processing Therapy; CPTS-RI = Child Post-Traumatic Stress - Reaction Index; CPTSDI = Child PTSD Inventory; CRI = Child Reaction Index; CRIES = Children's Revised Impact of Event Scale; CRTI = Children’s Responses to Trauma Inventory; CT-PTSD = Cognitive Therapy for PTSD; D-CPT = Developmentally adapted Cognitive Processing Therapy; DICA = Diagnostic Interview for Children and Adolescents; DISC = Diagnostic Interview Schedule for Children; DR Kongo = Democratic Republic of Kongo; EMDR = Eye Movement Desensitization and Reprocessing; EMDR-GP/C = EMDR-group protocol for children; EMDR-IGTP-OTS = EMDR-Integrative Group Treatment Protocol for Ongoing Traumatic Stress; ESGI = Expressive supportive group intervention; FAMOS = Family-Oriented Support; FU = Follow-Up; HTQ = Harvard Trauma Questionnaire; ITT = intent-to-treat data; K-SADS = Schedule for Affective Disorders and Schizophrenia for School-Age Children; KIDNET = Narrative Exposure Therapy for Children; MDT = multi-disciplinary treatment; MINI-KID = Mini International Neuropsychiatric Interview for Children and Adolescents; n.a. = not applicable; NET = Narrative Exposure Therapy; NL = the Netherlands; Non-TF-PI = non-trauma-focused psychological intervention; n.r. = not reported; NST = Non-directive Supportive Therapy; Other-TF-PI = other trauma-focused psychological intervention; PAPA = The Preschool Age Psychiatric Assessment; PCPI = Power up Children's Psychological Immunity; PE = Prolonged Exposure; PEDS = Pediatric Emotional Distress Scale; PDP = Psychodynamic Psychotherapy; PSS-SR = PTSD Symptom Scale, PST = Problem Solving Therapy; Self-Report; PTSD = post-traumatic stress disorder; PTSS-C = Posttraumatic Stress Symptoms Scale for Children; RRFT = Risk Reduction through Family Therapy; SC = Supportive Counselling; SF-AT = Solution-Focused Art Therapy; SHAT = Spiritual-Hypnosis Assisted Treatment; TARGET = Trauma Affect Regulation: Guide for Education and Therapy; TAU = Treatment as Usual; TF-CBT = trauma-focused cognitive behaviour therapy; TF-CBT-c = trauma-focused cognitive behaviour therapy with child only; TF-CBT-c&p = trauma-focused cognitive behaviour therapy with child and parent; TRT = Teaching Recovery Techniques; TRT+ = Teaching Recovery Techniques adapted version (to vicitims of natural disasters); TRT+P = Teaching Recovery Techniques plus parenting; TSCC = Trauma Symptom Checklist for Children; UC = usual care; UCLA PTSD-RI = University of California–Los Angeles PTSD Reaction Index; UK = United Kingdom; UPID = The University of California at Los Angeles PTSD Index; USA = United States of America; WL = Waitlist control condition (i.e., assessment only).

**Appendix G: References of studies included in the present network meta-analysis**

59.	Schauer E. *Trauma treatment for children in war: Build-up of an evidence-based large-scale mental health intervention in north-eastern Sri Lanka*. [Doctoral dissertation]; 2008.

**Appendix H: Trial and sample characteristics across comparison dyads**

|  | *Sample mean age* | | *% of participants identifying as female* | | *RCTs involving participants from high-income countries* | | *High-quality trials^a^* | | *PTSD outcome assessment with interview-based measure* | | *Trials with individual delivery of intervention^b^* | | *Treatment length in minutes^c^* | | *Parent / caregiver involvement in treatment = yes* | |
| --- | --- | --- | --- | --- | --- | --- | --- | --- | --- | --- | --- | --- | --- | --- | --- | --- |
| *Comparison* | *mean* | *SD* | *mean* | *SD* | *k* | *% of trials* | *k* | *% of trials* | *k* | *% of trials* | *k* | *% of trials* | *mean* | *SD* | *k* | *% of trials* |
| TF-CBTs vs PCC | 13.61 | 3.05 | 56.92 | 30.05 | 10 | 50 | 11 | 55 | 10 | 50 | 12 | 60 | 732.37 | 442.05 | 5 | 25 |
| EMDR vs PCC | 12.24 | 3.70 | 45.08 | 22.37 | 5 | 71.43 | 3 | 42.86 | 3 | 42.86 | 4 | 57.14 | 349.25 | 85.54 | 0 | 0 |
| non-TF-PIs vs PCC | n.r. | n.r. | 0 | 0 | 0 | 0 | 0 | 0 | 0 | 0 | 1 | 100 | 300 | 0 | 0 | 0 |
| MDTs vs PCC | 10.83 | 4.07 | 50.54 | 17.66 | 3 | 75 | 3 | 75 | 3 | 75 | 1 | 25 | 867.50 | 385.26 | 3 | 75 |
| TF-CBTs vs ACC | 13.14 | 3.35 | 69.29 | 21.41 | 9 | 60 | 8 | 53.33 | 9 | 60 | 10 | 66.67 | 626.96 | 251.97 | 7 | 46.47 |
| EMDR vs ACC | 9.60 | 0 | 33.78 | 0 | 1 | 100 | 1 | 100 | 0 | 0 | 1 | 100 | 175 | 0 | 1 | 100 |
| non-TF-PIs vs ACC | 3.45 | 0 | 59 | 0 | 1 | 100 | 1 | 100 | 0 | 0 | 0 | 0 | 525 | 0 | 1 | 100 |
| MDTs vs ACC | 14.13 | 0.84 | 87.72 | 17.50 | 3 | 75 | 1 | 25 | 4 | 100 | 2 | 50 | 1050 | 595.30 | 2 | 50 |
| TF-CBTs vs MDTs | 13.89 | 1.29 | 52.44 | 14.76 | 1 | 50 | 1 | 50 | 2 | 100 | 0 | 0 | 817.50 | 10.61 | 2 | 100 |
| TF-CBTs vs non-TF-PIs | 11.33 | 1.69 | 51.29 | 22.39 | 5 | 55.56 | 5 | 55.56 | 6 | 66.67 | 9 | 100 | 605.83 | 376.31 | 5 | 55.56 |
| TF-CBTs vs EMDR | 12.02 | 1.66 | 54.67 | 9.41 | 3 | 100 | 3 | 100 | 2 | 66.67 | 3 | 100 | 410 | 268.89 | 2 | 66.67 |

*Note.* Abbreviations (alphabetical): ACC = active control conditions; EMDR = Eye Movement Desensitization and Reprocessing; MDTs = multidisciplinary treatments; n.r. = (mean age) not reported; TF-CBTs = Trauma-Focused Cognitive Behavior Therapies (i.e., any CBT-based intervention with a trauma focus); PCC = Passive Control Conditions; non-TF-PIs = non-trauma-focused psychological interventions.
*^a^*Meeting at least six of eight trial quality criteria (see Appendices C and D above).
*^b^*Treatment delivered individually and face-to-face.
*^c^*Total treatment length in minutes ([mean] number of sessions multiplied by [mean] session length).

**Appendix I: Short-term efficacy: Forest plot compared to passive control conditions**

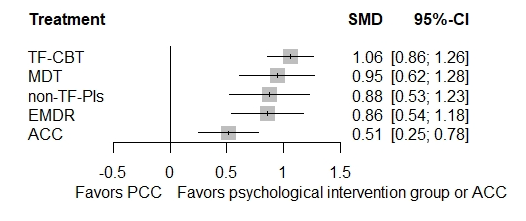

*Note*.	ACC = active control conditions (e.g., = treatment-as-usual); MDT = multi-disciplinary treatments (e.g., intensive multimodal group program); non-TF-PIs = non-trauma-focused psychological interventions (e.g., non-directive supportive therapy); PCC = passive control conditions (e.g., = waitlist); TF-CBT = trauma-focused cognitive behaviour therapies (e.g., prolonged exposure).

**Appendix J: Short-term efficacy: Forest plot compared to active control conditions**

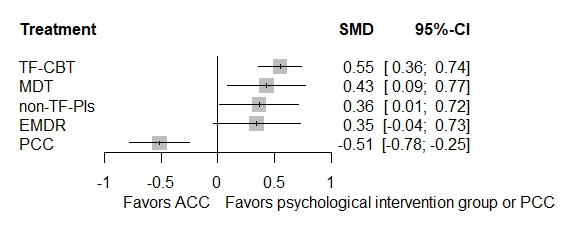

*Note*.	ACC = active control conditions (e.g., = treatment-as-usual); MDT = multi-disciplinary treatments (e.g., intensive multimodal group program); non-TF-PIs = non-trauma-focused psychological interventions (e.g., non-directive supportive therapy); PCC = passive control conditions (e.g., = waitlist); TF-CBT = trauma-focused cognitive behaviour therapies (e.g., prolonged exposure).

**Appendix K: Short-term efficacy: Net splitting results**

| comparison dyads | kes | prop | nma | direct effect | indirect effect | Difference in effects | z | p-value |
| --- | --- | --- | --- | --- | --- | --- | --- | --- |
| ACC vs. EMDR | 0 | 0 | -0.3450 | n.a. | -0.3450 | n.a. | n.a. | n.a. |
| ACC vs. MDTs | 4 | 0.47 | -0.4314 | -0.3347 | -0.5174 | 0.1828 | 0.52 | 0.5999 |
| ACC vs. non-TF-PIs | 0 | 0 | -0.3643 | n.a. | -0.3643 | n.a. | n.a. | n.a. |
| ACC vs. PCC | 1 | 0.06 | 0.5147 | 0.1800 | 0.5343 | -0.3543 | -0.61 | 0.5438 |
| ACC vs. TF-CBTs | 21 | 0.92 | -0.5494 | -0.5427 | -0.6301 | 0.0874 | 0.24 | 0.8069 |
| EMDR vs. MDTs | 0 | 0 | -0.0863 | n.a. | -0.0863 | n.a. | n.a. | n.a. |
| EMDR vs. non-TF-PIs | 0 | 0 | -0.0193 | n.a. | -0.0193 | n.a. | n.a. | n.a. |
| EMDR vs. PCC | 7 | 0.74 | 0.8597 | 0.7776 | 1.0889 | -0.3113 | -0.84 | 0.4030 |
| EMDR vs. TF-CBTs | 3 | 0.44 | -0.2043 | 0.0556 | -0.4097 | 0.4653 | 1.33 | 0.1843 |
| MDTs vs. non-TF-PIs | 0 | 0 | 0.0671 | n.a. | 0.0671 | n.a. | n.a. | n.a. |
| MDTs vs. PCC | 4 | 0.55 | 0.9461 | 0.9329 | 0.9622 | -0.0293 | -0.09 | 0.9311 |
| MDTs vs. TF-CBTs | 2 | 0.22 | -0.1180 | 0.0949 | -0.1778 | 0.2727 | 0.67 | 0.5038 |
| non-TF-PIs vs. PCC | 1 | 0.13 | 0.8790 | 1.2999 | 0.8187 | 0.4812 | 0.88 | 0.3763 |
| non-TF-PIs vs. TF-CBTs | 9 | 0.97 | -0.1850 | -0.1808 | -0.3424 | 0.1616 | 0.17 | 0.8657 |
| TF-CBTs vs. PCC | 18 | 0.80 | 1.0640 | 1.0898 | 0.9581 | 0.1317 | 0.51 | 0.6097 |

*Note*.	ACC = active control conditions (e.g., = treatment-as-usual); EMDR = Eye Movement Desensitization and Reprocessing; MDTs = multi-disciplinary treatments (e.g., intensive multimodal group program); non-TF-PIs = non-trauma-focused psychological interventions (e.g., non-directive supportive therapy); PCC = passive control conditions (e.g., = waitlist); TF-CBTs = trauma-focused cognitive behaviour therapies (e.g., prolonged exposure).

**Appendix L: Short-term efficacy: Net heat plot**

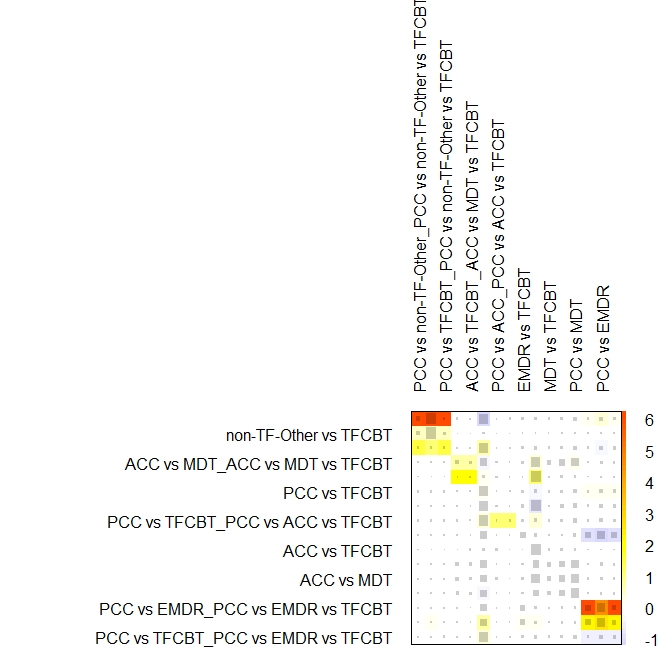

*Note*.	ACC = active control conditions (e.g., = treatment-as-usual); MDT = multi-disciplinary treatments (e.g., intensive multimodal group program); non-TF-Other = non-trauma-focused psychological interventions (e.g., non-directive supportive therapy); PCC = passive control conditions (e.g., = waitlist); TF-CBT = trauma-focused cognitive behaviour therapies (e.g., prolonged exposure).

**Appendix M: Short-term efficacy: Funnel plot**

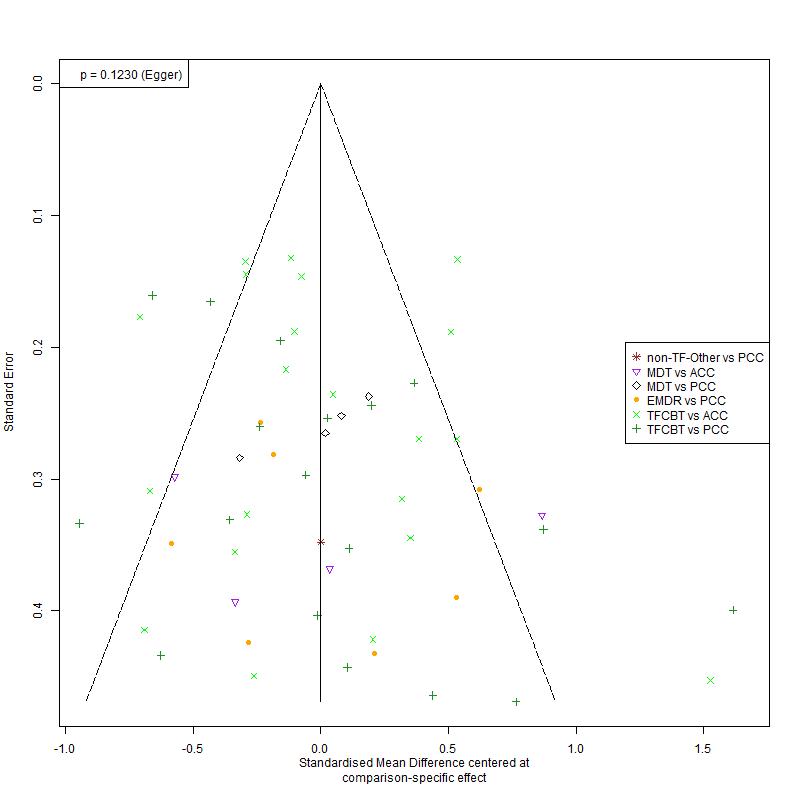

*Note*.	ACC = active control conditions (e.g., = treatment-as-usual); MDT = multi-disciplinary treatments (e.g., intensive multimodal group program); non-TF-Other = non-trauma-focused psychological interventions (e.g., non-directive supportive therapy); PCC = passive control conditions (e.g., = waitlist); TF-CBT = trauma-focused cognitive behaviour therapies (e.g., prolonged exposure).

**Appendix N: Short-term efficacy: Outlier-adjusted results**

| *Reference group* | | | *Psych. interv.* |  | *kes (N)* | *SMD* | *95% CI* | *p* | *I^2^ (τ^2^)* |
| --- | --- | --- | --- | --- | --- | --- | --- | --- | --- |
|  |  | ***Main analysis (i.e., irrespective of trial quality and delivery format)*** | | | | | | |  |
| relative to PCC | | TF-CBTs# | |  | 17 (1,097) | **0.99** | **[0.80 – 1.19]** | **< .001** | 64.2  ***  (0.11) |
|  |  | EMDR | |  | 7 (297) | **0.84** | **[0.54 – 1.14]** | **< .001** |  |
|  |  | non-TF-PIs | |  | 1 (40) | **0.80** | **[0.47 – 1.13]** | **< .001** |  |
|  |  | MDTs | |  | 4 (270) | **0.93** | **[0.62 – 1.23]** | **< .001** |  |
|  | | ACC | |  | 1 (22) | **0.49** | **[0.25 – 0.74]** | **<.001** |  |
| relative to ACC | | TF-CBTs# | |  | 20 (2,018) | **0.50** | **[0.33 – 0.68]** | **< .001** |  |
|  |  | EMDR | |  | 0 (0) | 0.35 | [-0.01 – 0.70] | .059 |  |
|  |  | non-TF-PIs | |  | 0 (0) | 0.31 | [-0.01 – 0.64] | .061 |  |
|  |  | MDTs | |  | 4 (146) | **0.43** | **[0.12 – 0.75]** | **.008** |  |
| relative to EMDR | | TF-CBTs | |  | 3 (185) | 0.16 | [-0.16 – 0.48] | .334 |  |
|  |  | non-TF-PIs | |  | 0 (0) | -0.03 | [-0.45 – 0.39] | .877 |  |
|  |  | MDTs | |  | 0 (0) | 0.09 | [-0.33 – 0.51] | .673 |  |
| relative to  non-TF-PIs | | TF-CBTs | |  | 9 (631) | 0.19 | [-0.09 – 0.47] | .177 |  |
|  |  | MDTs | |  | 0 (0) | 0.12 | [-0.29 – 0.54] | .560 |  |
| relative to MDTs | | TF-CBTs | |  | 2 (72) | 0.07 | [-0.24 – 0.38] | .668 |  |

*Note*.	ACC = active control conditions (e.g. = treatment-as-usual); EMDR = eye movement desensitization and reprocessing; kes = number of direct comparisons for the given comparison; MDTs = multidisciplinary treatments; N = total number of participants; non-TF-PIs = non-trauma-focused psychological interventions; PCC = passive control conditions (e.g. = waitlist); Psych. interv. = psychological interventions; SMD = standardized mean differences (i.e. = Hedges’ g); TF-CBTs = trauma-focused cognitive behaviour therapies. **Bold** font highlights statistical significance of findings. A positive (negative) SMD indicates superior (inferior) efficacy of the given psychological intervention relative to the given reference group.
*** p < .001, ** p < .01, * p < .05, corresponding to the respective Q-statistic as a measure of heterogeneity in outcomes.

\# 2 statistical outliers (i.e., McMullen et al., 2013 comparing TF-CBT to PCC & Barron et al., 2021 comparing TF-CBT to ACC) were excluded from this outlier-adjusted analyses.

**Appendix O: Mid-term efficacy: Forest plot compared to passive control conditions**

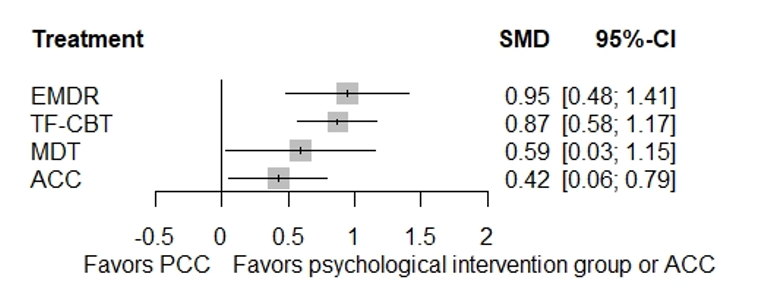

*Note*.	ACC = active control conditions (e.g., = treatment-as-usual); EMDR = eye movement desensitization and reprocessing; MDT = multi-disciplinary treatments (e.g., intensive multimodal group program); PCC = passive control conditions (e.g., = waitlist); TF-CBT = trauma-focused cognitive behaviour therapies (e.g., prolonged exposure).

**Appendix P: Mid-term efficacy: Forest plot compared to active control conditions**

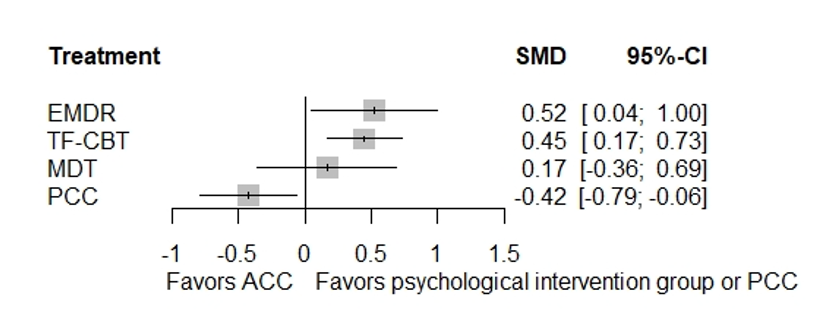

*Note*.	ACC = active control conditions (e.g., = treatment-as-usual); EMDR = eye movement desensitization and reprocessing; MDT = multi-disciplinary treatments (e.g., intensive multimodal group program); PCC = passive control conditions (e.g., = waitlist); TF-CBT = trauma-focused cognitive behaviour therapies (e.g., prolonged exposure).

**Appendix Q: Mid-term efficacy: Funnel plot**

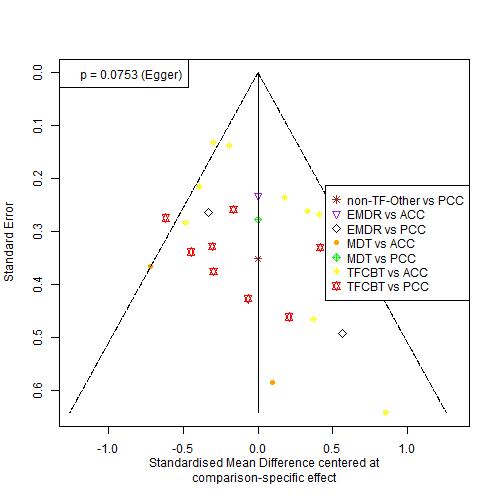

*Note*.	ACC = active control conditions (e.g., = treatment-as-usual); EMDR = eye movement desensitization and reprocessing; MDT = multi-disciplinary treatments (e.g., intensive multimodal group program); PCC = passive control conditions (e.g., = waitlist); TF-CBT = trauma-focused cognitive behaviour therapies (e.g., prolonged exposure).

**Appendix R: Mid-term efficacy: Net splitting results**

| comparison dyads | kes | prop | nma | direct effect | indirect effect | Difference in effects | z | p-value |
| --- | --- | --- | --- | --- | --- | --- | --- | --- |
| ACC vs. EMDR | 1 | 0.30 | -0.5212 | 0.0386 | -0.7570 | 0.7956 | 1.49 | 0.1358 |
| **ACC vs. MDTs** | **3** | **0.66** | **-0.1664** | **-0.5535** | **0.5995** | **-1.1530** | **-2.03** | **0.0424** |
| ACC vs. PCC | 2 | 0.26 | 0.4240 | 0.2602 | 0.4827 | -0.2225 | -0.52 | 0.6016 |
| ACC vs. TF-CBTs | 9 | 0.85 | -0.4501 | -0.4241 | -0.5938 | 0.1697 | 0.42 | 0.6724 |
| EMDR vs. MDTs | 0 | n.a. | 0.3548 | n.a. | 0.3548 | n.a. | n.a. | n.a. |
| EMDR vs. PCC | 2 | 0.41 | 0.9451 | 1.1923 | 0.7746 | 0.4177 | 0.86 | 0.3883 |
| EMDR vs. TF-CBTs | 2 | 0.47 | 0.0711 | 0.1812 | -0.0270 | 0.2081 | 0.46 | 0.6458 |
| MDTs vs. PCC | 1 | 0.37 | 0.5903 | -0.0950 | 0.9884 | -1.0834 | -1.82 | 0.0682 |
| MDTs vs. TF-CBTs | 1 | 0.19 | -0.2837 | -0.3879 | -0.2600 | -0.1280 | -0.18 | 0.8584 |
| TF-CBTs vs. PCC | 9 | 0.80 | 0.8741 | 0.9187 | 0.6989 | 0.2198 | 0.58 | 0.5603 |

*Note*.	ACC = active control conditions (e.g., = treatment-as-usual); EMDR = Eye Movement Desensitization and Reprocessing; MDTs = multi-disciplinary treatments (e.g., intensive multimodal group program); PCC = passive control conditions (e.g., = waitlist); TF-CBTs = trauma-focused cognitive behaviour therapies (e.g., prolonged exposure).

**Appendix S: Mid-term efficacy: Net heat plot before (top) and after (bottom) exclusion of MDTs**

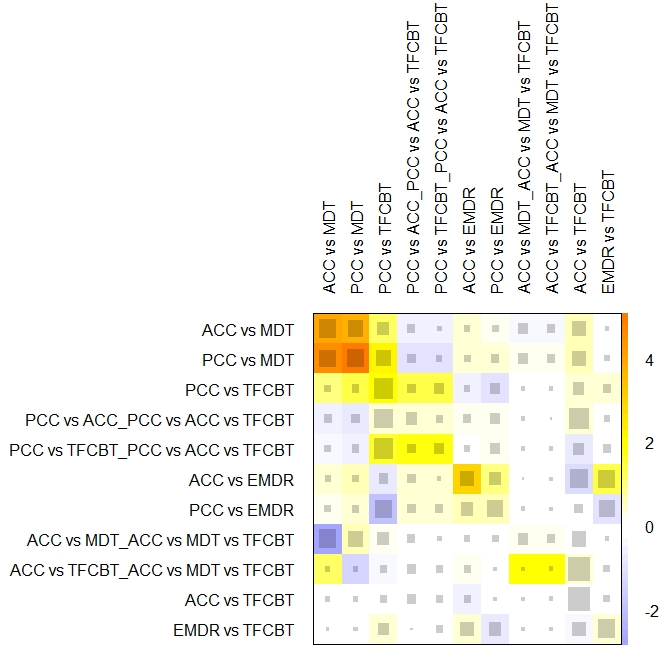

Note.	ACC = active control conditions (e.g., = treatment-as-usual); EMDR = eye movement desensitization and reprocessing; MDT(s) = multi-disciplinary treatments (e.g., intensive multimodal group program); PCC = passive control conditions (e.g., = waitlist); TF-CBT = trauma-focused cognitive behaviour therapies (e.g., prolonged exposure).

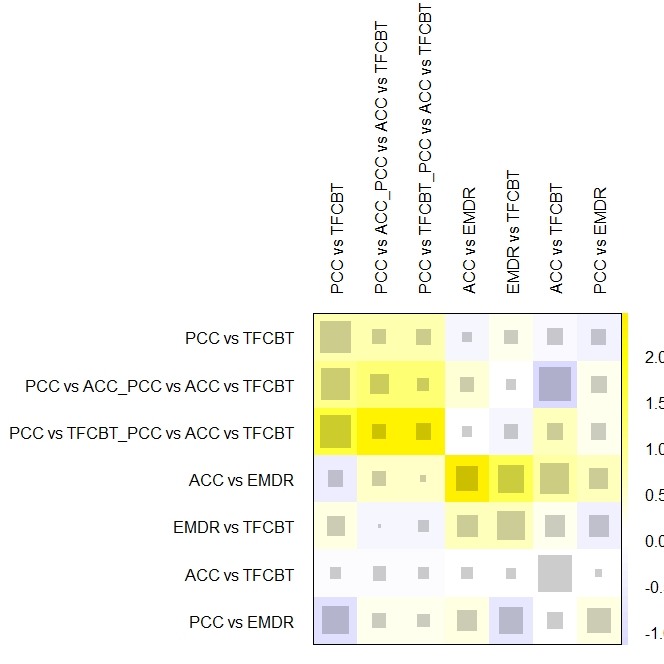

Note.	ACC = active control conditions (e.g., = treatment-as-usual); EMDR = eye movement desensitization and reprocessing; MDT = multi-disciplinary treatments (e.g., intensive multimodal group program); PCC = passive control conditions (e.g., = waitlist); TF-CBT = trauma-focused cognitive behaviour therapies (e.g., prolonged exposure).

**Appendix T: Mid-term efficacy: MDTs excluded due to detected inconsistency**

| *Reference group Psych. interv.* | | | *kes (N)* | *SMD* | *95% CI* | *p* | *I2 (τ2)* |
| --- | --- | --- | --- | --- | --- | --- | --- |
|  | **FU1 (i.e., ≤ 5 months follow-up) *-* main analysis** | | | | | |  |
| relative to PCC | | TF-CBTs | 9 (389) | **0.94** | **[0.65 – 1.22]** | **< .001** | 62.9  ***  (0.11) |
|  |  | EMDR | 2 (86) | **0.99** | **[0.56 – 1.42]** | **< .001** |  |
|  | | ACC | 2 (74) | **0.54** | **[0.19 – 0.90]** | **.003** |  |
| relative to ACC | | TF-CBTs | 9 (813) | **0.39** | **[0.13 – 0.66]** | **.003** |  |
|  |  | EMDR | 1 (74) | **0.45** | **[0.01 – 0.89]** | **.046** |  |
| relative to EMDR | | TF-CBTs | 2 (125) | -0.05 | [-0.46 – 0.35] | .799 |  |

*Note*.	ACC = active control conditions (e.g. = treatment-as-usual); EMDR = eye movement desensitization and reprocessing; kes = number of direct comparisons for the given comparison; MDTs = multidisciplinary treatments; N = total number of participants; PCC = passive control conditions (e.g. = waitlist); Psych. interv. = psychological interventions; SMD = standardized mean differences (i.e. = Hedges’ g); TF-CBTs = trauma-focused cognitive behaviour therapies.
**Bold** font highlights statistical significance of findings. A positive (negative) SMD indicates superior (inferior) efficacy of the given psychological intervention relative to the given reference group.
*** p < .001, ** p < .01, * p < .05, corresponding to the respective Q-statistic as a measure of heterogeneity in outcomes.

**Appendix U: Long-term efficacy: Forest plot compared to passive control conditions**

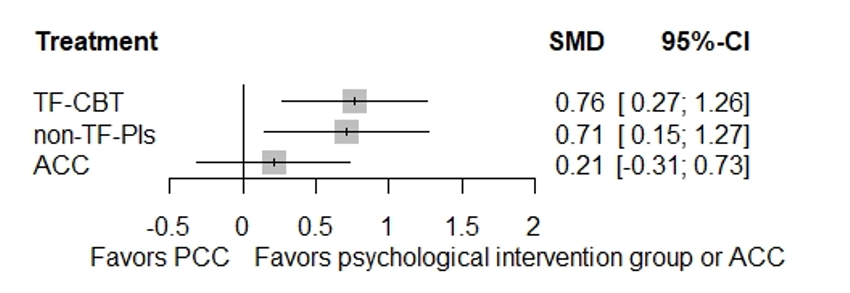

*Note*.	ACC = active control conditions (e.g., = treatment-as-usual); non-TF-PIs = non-trauma-focused psychological interventions (e.g., non-directive supportive therapy); PCC = passive control conditions (e.g., = waitlist); TF-CBTs = trauma-focused cognitive behaviour therapies (e.g., prolonged exposure).

**Appendix V: Long-term efficacy: Forest plot compared to active control conditions**

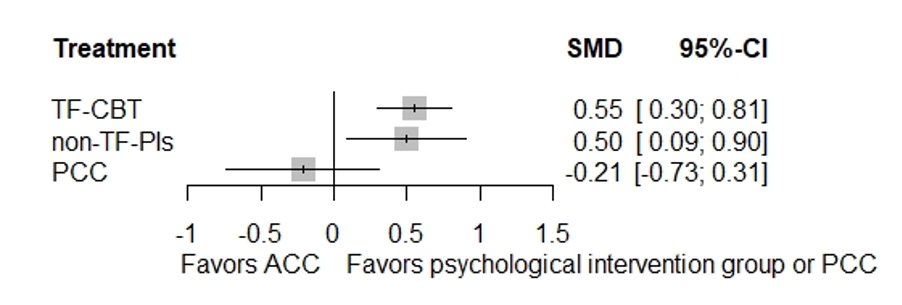

*Note*.	ACC = active control conditions (e.g., = treatment-as-usual); non-TF-PIs = non-trauma-focused psychological interventions (e.g., non-directive supportive therapy); PCC = passive control conditions (e.g., = waitlist); TF-CBTs = trauma-focused cognitive behaviour therapies (e.g., prolonged exposure).

**Appendix W: Long-term efficacy: Net splitting results**

| comparison dyads | kes | prop | nma | direct effect | indirect effect | Difference in effects | z | p-value |
| --- | --- | --- | --- | --- | --- | --- | --- | --- |
| ACC vs. non-TF-PIs | 1 | 0.20 | -0.4982 | -0.8293 | -0.4140 | -0.4153 | -0.81 | 0.4203 |
| ACC vs. PCC | 1 | 0.37 | 0.2107 | 0.0354 | 0.3147 | -0.2793 | -0.50 | 0.6137 |
| ACC vs. TF-CBTs | 9 | 0.92 | -0.5539 | -0.5091 | -1.0608 | 0.5516 | 1.16 | 0.2451 |
| non-TF-PIs vs. PCC | 1 | 0.36 | 0.7089 | 1.2387 | 0.4162 | 0.8225 | 1.37 | 0.1706 |
| non-TF-PIs vs. TF-CBTs | 5 | 0.84 | -0.0557 | -0.1320 | 0.3404 | -0.4723 | -0.98 | 0.3287 |
| TF-CBTs vs. PCC | 3 | 0.83 | 0.7646 | 0.6403 | 1.3929 | -0.7526 | -1.11 | 0.2649 |

*Note*.	ACC = active control conditions (e.g., = treatment-as-usual); non-TF-PIs = non-trauma-focused psychological interventions (e.g., non-directive supportive therapy); PCC = passive control conditions (e.g., = waitlist); TF-CBTs = trauma-focused cognitive behaviour therapies (e.g., prolonged exposure).

**Appendix X: Long-term efficacy: Net heat plot**

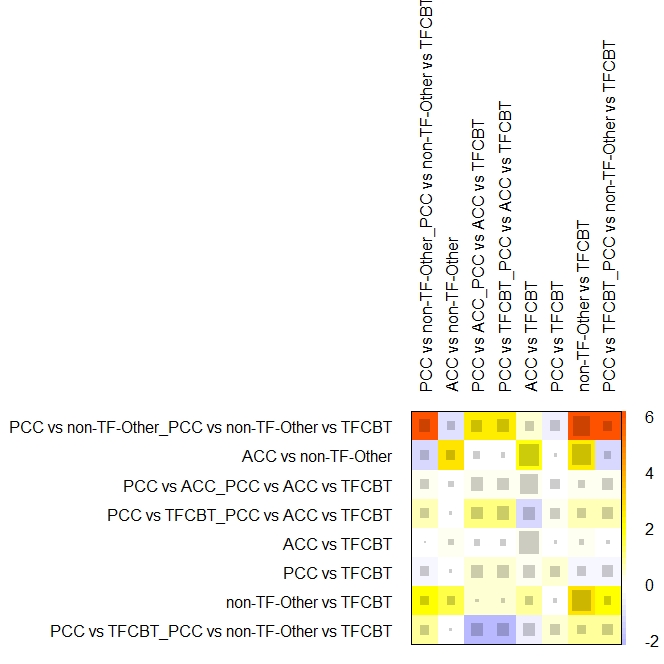

*Note*.	ACC = active control conditions (e.g., = treatment-as-usual); non-TF-Other = non-trauma-focused psychological interventions (e.g., non-directive supportive therapy); PCC = passive control conditions (e.g., = waitlist); TF-CBTs = trauma-focused cognitive behaviour therapies (e.g., prolonged exposure).

**Appendix Y: Long-term efficacy: Funnel plot**

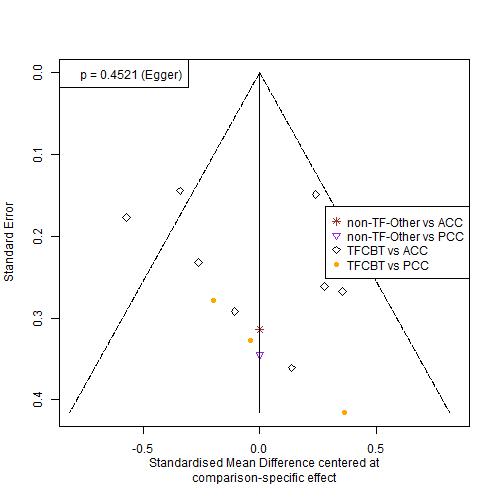

*Note*.	ACC = active control conditions (e.g., = treatment-as-usual); non-TF-Other = non-trauma-focused psychological interventions (e.g., non-directive supportive therapy); PCC = passive control conditions (e.g., = waitlist); TF-CBTs = trauma-focused cognitive behaviour therapies (e.g., prolonged exposure).
